# A highly prevalent lupus risk haplotype increases IRF7-dependent induction of IFN-α, enhancing antiviral defense and exacerbating autoimmunity

**DOI:** 10.64898/2026.01.21.26344474

**Authors:** Samuel J. Virolainen, Kathryn Creighton, Maryam Dashtiahangar, Durga Krishnamurthy, Lois Parks, Carmy Forney, Britney Ampadu, Akshata N. Rudrapatna, Katelyn A. Dunn, Sreeja Parameswaran, Hayley K. Hesse, Xiaoting Chen, Andrew VonHandorf, Lee E. Edsall, Cailing Yin, Arthur Lynch, Olivia E. Gittens, Arame A. Diouf, Sydney H. Jones, Matthew Hass, Ellen Javier, Omer A. Donmez, Yasine Keddari, Oded Danzinger, Harsha Seelamneni, Bahram Namjou-Khales, Hannah C. Ainsworth, Mary E. Comeau, Miranda C. Marion, Stuart B. Glenn, Swapan K. Nath, Barry I. Freedman, Betty P. Tsao, Diane L. Kamen, Elizabeth E. Brown, Gary S. Gilkeson, Graciela S. Alarcón, John D. Reveille, Judith A. James, Lindsey A. Criswell, Luis M. Vilá, Marta E. Alarcón-Riquelme, Michelle Petri, R. Hal Scofield, Robert P. Kimberly, Rosalind Ramsey-Goldman, Sang-Cheol Bae, Deborah Cunninghame Graham, Timothy J. Vyse, Joel M. Guthridge, Patrick M. Gaffney, Carl D. Langefeld, Jennifer A. Kelly, Kenneth M. Kaufman, Kathy Sivils, John B. Harley, Nan Shen, Lucinda P. Lawson, Yuriy Baglaenko, Emily R. Miraldi, Brad R. Rosenberg, Trevor Siggers, Stephen N. Waggoner, Matthew T. Weirauch, Leah C. Kottyan

## Abstract

Genome-wide association studies have identified genetic polymorphisms at 11p15 associated with Systemic Lupus Erythematosus (lupus). Statistical fine mapping prioritizes a highly prevalent coding haplotype within the IRF7 gene. Analysis of ancient DNA confirms that this haplotype has persisted at high frequencies in the global population for millennia. The IRF7 risk haplotype is sufficient to increase nuclear localization of IRF7 and transcriptional activity downstream of pattern recognition receptor pathways. This risk haplotype increases IRF7 DNA binding strength and alters IRF7 DNA sequence specificity, resulting in genotype-dependent increases in IFN-α production in numerous biological systems, including monocytes and airway epithelial cells. CRISPR engineering of a homologous risk variant in mouse Irf7 results in both enhanced innate control of virus infection and increased autoantibody titers in a model of autoimmunity. Altogether, we establish a persistent and prominent genetic IRF7 haplotype that amplifies IRF7 activity in a manner that has immunological risks and benefits.

**HIGHLIGHTS:**

● Genetic analysis using modern and evolutionary datasets identifies a persistent and highly prevalent lupus-associated coding haplotype in *IRF7* at 11p15
● The *IRF7* lupus risk haplotype increases IFN-α production by monocytes and airway epithelial cells
● The *IRF7* lupus risk haplotype increases IRF7 DNA binding strength and alters DNA sequence specificity
● A homologous lupus risk variant in mouse *Irf7* enhances control of vesicular stomatitis virus and exacerbates autoantibody production

## INTRODUCTION

Systemic lupus erythematosus (SLE or lupus) is a debilitating autoimmune disease driven by widespread inflammation, immune complex formation, and autoantibody production ^1,2^. The heritability of lupus is between 43 and 66% across ancestral groups, and over 200 genetic risk loci have been associated with lupus through genome wide association studies (GWAS) ^3–10^. Genetic risk loci for lupus are highly enriched for type I interferon (IFN-I) signaling pathway components ^11,12^. IFN-I production is initiated upon sensing of viruses by pattern recognition receptors (PRRs), leading to an inflammatory signaling cascade and a gene expression program that restricts viral growth and influences innate and adaptive immune responses ^13^. Thus, potent IFN-I responses are critical to clear viral infections, but dysregulation of this pathway contributes to autoimmunity and lupus ^14^.

Interferon regulatory factor-7 (IRF7) is sometimes referred to as the master regulator of the IFN-I response ^13,15^. IRF7 is activated downstream of pattern recognition receptors (PRRs) ^15–19^ that include Toll-like receptors (TLRs), retinoic acid-inducible gene I (RIG-I), and cyclic GMP-AMP synthase (cGAS)-stimulator of interferon genes (STING). IRF7 activation leads to its nuclear localization and binding to DNA, with subsequent induction of interferon genes and IFN-I secretion ^16,18,20–22^. Secreted IFN-I binds the IFN-I receptor (IFNAR) to initiate autocrine and paracrine signaling cascades that restrict virus replication, induce interferon stimulated genes (ISGs), propagate inflammatory responses, and contribute to adaptive immune responses ^13,23^. Robust IRF7 driven IFN-I responses are crucial to human health, as IRF7 deficiency and autoantibodies that neutralize IFN-I each increase the risk of hospitalization and death from viruses such as influenza virus A (IAV) and SARS-CoV-2 ^24–27^.

IRF7-driven IFN-I responses also contribute to pathogenesis of autoimmune diseases, including lupus ^28,29^. More than 50% of patients with lupus have elevated IFN-I in peripheral blood, a phenomenon known as the “IFN-I Signature” of lupus ^30^. In addition, autoantibody production in mouse models of lupus is *Irf7* dependent ^31^. Variants at the chromosome 11p15 locus encoding *IRF7* are associated with differential risk for lupus development ^32–35^. Specifically, a human genetic variant (rs1131665) results in a coding amino acid change from R to Q at position 412 of the IRF7 protein, which is the most robustly associated variant at the *IRF7* locus across numerous studies ^32,33,36^. Further, this variant is highly prevalent in the global population, with major allele frequencies approaching nearly 98% in certain populations ^32,33^. However, much remains to be explored about the biological role of genetic variation at the *IRF7* locus and its impact on lupus risk. As IRF7 is a critical regulator of both antiviral and autoimmune-related IFN-I responses, and IRF7 genetic risk alleles are highly abundant in the human population, an improved understanding of the precise causal genetic variants and mechanisms of action is necessary to inform the development of new therapies and response to treatment in these important health contexts.

In this study, we explore the evolutionary persistence of this highly prevalent lupus-associated haplotype in *IRF7* (Q412R: K179E) and its relationship with the magnitude of IFN-I responses. Ancient DNA datasets are combined with viral challenge experiments to test the hypothesis that coding variants in *IRF7* persisted across human evolution based on their ability to enhance IFN-mediated antiviral immunity, with increased risk of lupus being an evolutionary consequence of this benefit to host defense. Mechanistically, we explore the effect of *IRF7* coding risk variants on IRF7 nuclear translocation, interactions with DNA, transcriptional regulatory activity, and IFN-α production downstream of innate pattern recognition receptors. Altogether, we establish a persistent and prominent genetic *IRF7* haplotype that amplifies IRF7 activity in a manner that has immunological risks and benefits.

## RESULTS

### Highly prevalent coding variants account for lupus risk at the *IRF7* locus

Genetic variants at the 11p15 locus are associated with lupus risk across ancestries ^10,32,33^. To determine the likely causal variant(s) driving the association signal at this locus, we interrogated a subset of the Large Lupus Association Study 2 that includes 150 genotyped variants at the 11p15 locus in a multi-ancestral cohort of 7,137 individuals with lupus and 6,329 individuals without lupus ^37^. We first stratified the population into four groups based on ancestrally informative markers and applied imputation to incorporate variants that were not genotyped (see **Methods**). We then performed an association study in European and African ancestries that conferred 96% and 80% power, respectively, to identify the variants in the 11p15 locus underlying risk for lupus (see **Table S1** and **Methods**). Analysis of other ancestral groups had insufficient power (<40%) due to the combined effects of lower minor allele prevalence and a small cohort size. We employed a logistic regression model with admixture estimates as covariates to identify highly associated variants **(Figure 1)**. In the European ancestry population, a haplotype of 17 variants in strong linkage disequilibrium (LD) exceeded the statistical threshold of genome-wide significance **(Figure 1A**, **Table 1).** This same haplotype reached modest (but not genome-wide) significance in the population of individuals with African ancestry **(Figure 1B**, **Table 1**). This haplotype achieved a P-value of 3.58×10^−10^ in a meta-analysis of the European and African ancestry populations **(Figure 1C**, **Table 1).** Two coding variants, rs1131665 (Q412R) and rs1061502 (K179E), were found within this robustly associated haplotype **(Figure 1D-E)**. The additive effect sizes of 1.15-1.20 **(Table 1)** for these two variants are consistent with prior studies ^32,33^. In contrast to most autoimmune-associated variants ^11^, the lupus risk haplotype for these variants is highly prevalent in multiple human populations, ranging from 70 to 98% across ancestries (**Figure 1F**).

**Figure 1.**
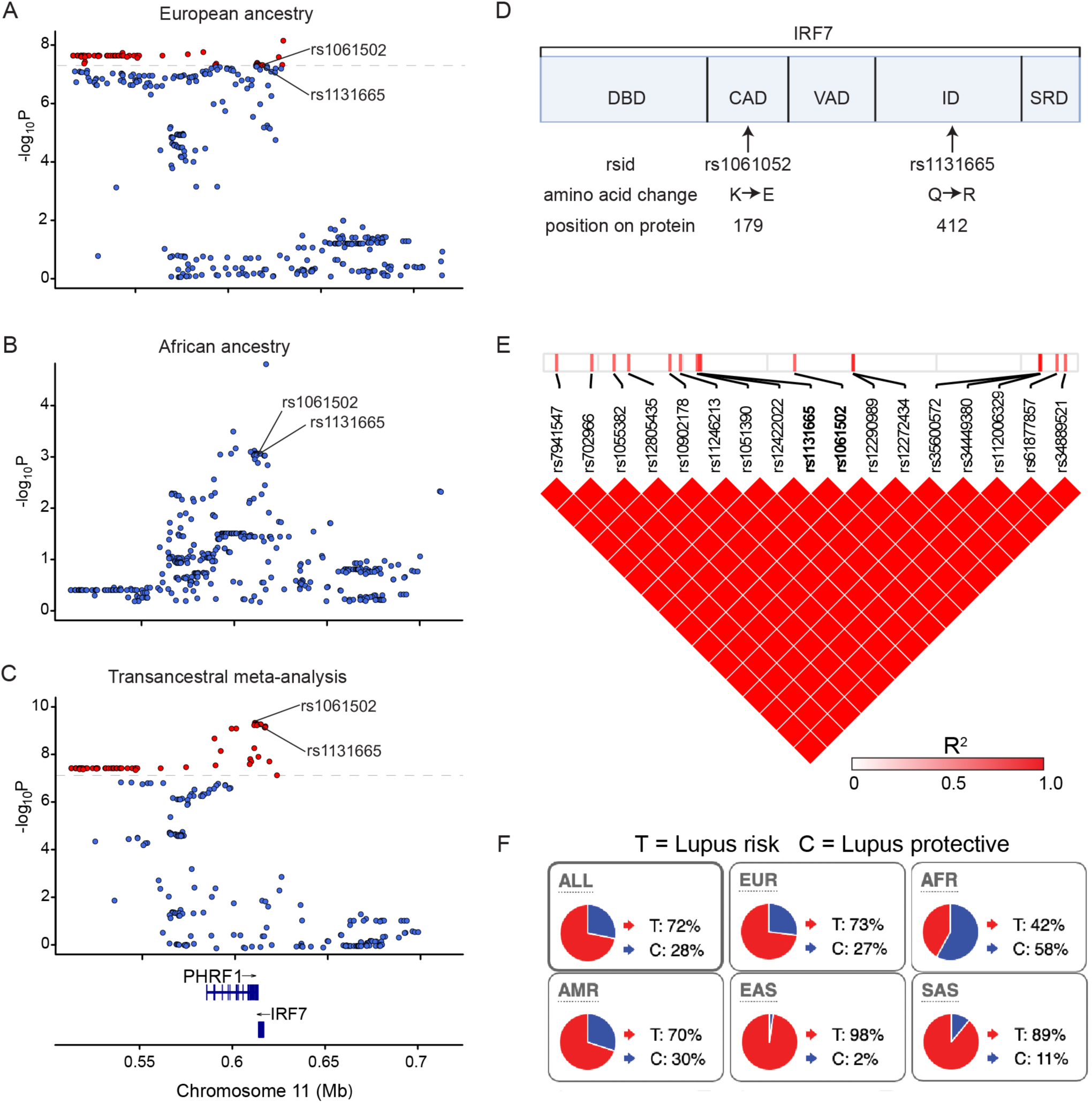
A highly prevalent haplotype in *IRF7* tagged by rs1131665 (Q412R) and rs1061502 (K179E) shows genome-wide association with lupus. (A-C) Genome-wide association signal of genetic variants in the *IRF7-PHRF1* locus. Each variant is represented as a data point in the context of its genomic location within the locus and is colored based on its P-value, with red dots representing genome-wide significant genetic variants (P<5×10-8) and blue dots signifying variants that do not meet this threshold. rs1131665 and rs1061502, which are characterized in this study, are identified in A-C. The variants were assessed in a logistic regression model using admixture estimates as a covariate. Genomic position is provided using GRCh37 (hg19) coordinates. (A) European Ancestry, (B) African Ancestry, (C) Trans-ancestral Meta-Analysis. (D) Schematic of IRF7 (isoform A) protein domains: DBD: DNA Binding Domain; CAD: Constitutive Activation Domain; VAD: Virus-Activated Domain; ID: Inhibitory Domain; SRD: Signal Response Domain. (E) Heatmap showing variants in the IRF7 haplotype assessed in the trans-ancestral meta-analysis demonstrating perfect linkage disequilibrium of *IRF7-PHRF1* variants; (F) Pie charts quantifying the frequency of the lupus risk (“T”) and protective (“C”) haplotypes globally and in specified ancestries (Ensembl database). Ancestry abbreviations (defined by Ensembl Gene): ALL = Global Population; EUR = European Ancestry; AFR = African Ancestry; AMR = American Ancestry (native people of the Americas); EAS = East Asian Ancestry; SAS = South Asian Ancestry.

**Table 1.**
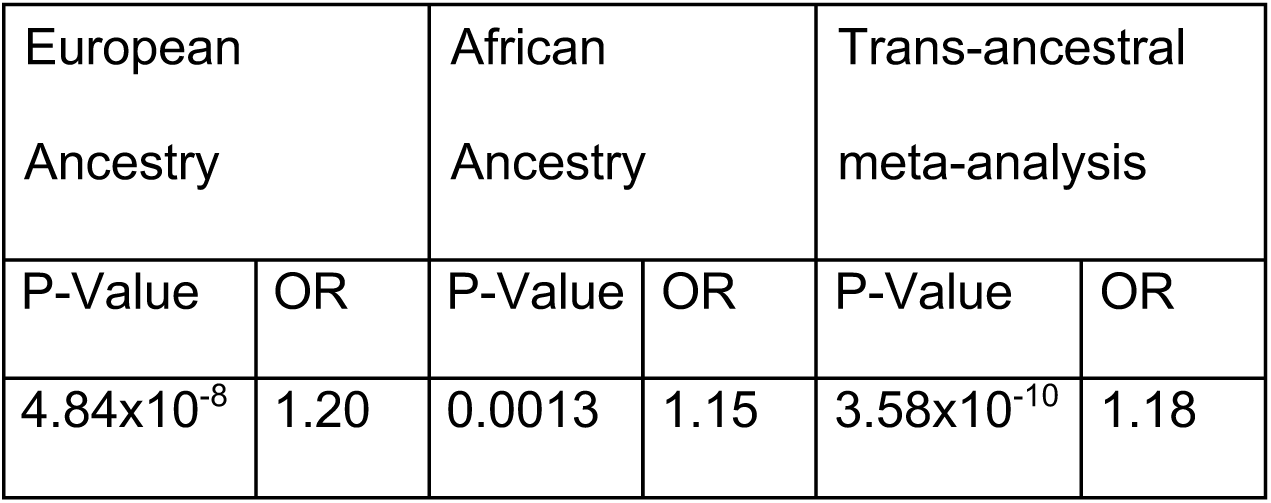
Lupus association for the *IRF7* haplotype block containing rs1131665 and rs1061502. P-values are based on a logistic regression model performed in each ancestry with admixture estimates as covariates. Odds ratios are calculated in terms of the risk haplotype. The trans-ancestral meta-analysis was performed using an inverse-variance model using data from the European and African cohorts only. Identical results were obtained when each variant in the haplotype was assessed individually because of the perfect LD (r^2^=1) between the variants. LD: Linkage Disequilibrium; OR: Odds Ratio.

To assess whether the haplotype containing Q412R (rs1131665) and K179E (rs1061502) accounts for lupus risk at 11p15, we performed a stepwise logistic regression analysis starting with the genotype of rs1131665 as a covariate in the cohort of European ancestry. The genotype of rs1131665 was sufficient to account for the entire association with a significance level of P<0.01 at this locus in the European ancestry cohort **(Figure S1A).** We next performed a separate stepwise logistic regression including rs1061502 as a covariate. As expected, the genotype of rs1061502 was also sufficient to account for the lupus association (P<0.01) at this locus **(Figure S1B).** This association is consistent with the inheritance of these two coding variants in a haplotype given their complete linkage disequilibrium (LD) (r^2^=1.0, **Figure 1E)**. This finding is especially important given that this risk haplotype is the most prevalent haplotype globally **(Figure 1F).** Collectively, our statistical analyses support a model in which lupus risk at the 11p15 (*IRF7*) locus is driven by the rs1131665+rs1061502 (*IRF7* Q412R+K179E) haplotype. In this study, we will refer to the Q412+K179 haplotype as “lupus risk” and the R412+E179 haplotype as “lupus protective.”

### A common lupus *IRF7* risk haplotype is highly conserved in ancient DNA

The high prevalence of the lupus-associated haplotype in multiple ancestries is striking, particularly because the frequency of disease-associated variants is often kept low by natural selection ^38–41^. Ancient DNA studies assessing rapidly changing allele frequencies in populations before and after pandemics or plagues can be used to reveal selection of immune-related genes^42^. To assess the conservation of this haplotype and determine potential selection events, we performed an analysis of publicly available datasets of ancient human DNA from the Ancient Genome Diversity Project – Allen Ancient DNA Resource (AADR) (see **Methods**) ^43^. We used rs11246213 as a tag for the rs1131665+rs106152 haplotype in this analysis. This dataset has global sampling **(Figure 2A),** with genomes as old as ∼100,000 before present. As this dataset represents only ∼1.23 million positions in the genome, our two lead SNPs were not represented. However, one of the 11p15 SNPs in LD with our two lead SNPs (perfect LD (R^2^=1) in EUR and AFR ancestry and nearly perfect (R^2^=0.95) in East Asian ancestry) was present in this dataset (rs11246213). Thus, this SNP was used to track population frequencies of this haplotype through time. Across the timeframe best captured in this dataset (∼10,000 BCE to ∼1,800 CE), the haplotype frequencies of the lupus risk allele remained largely consistent across ancestries, with high frequencies of the risk allele, and did not appear to respond to the increased immune threats (e.g., Black Death) associated with growing populations through time (first documented epidemics and pandemics, ∼165 CE onward) **(Figure 2B)**.

**Figure 2.**
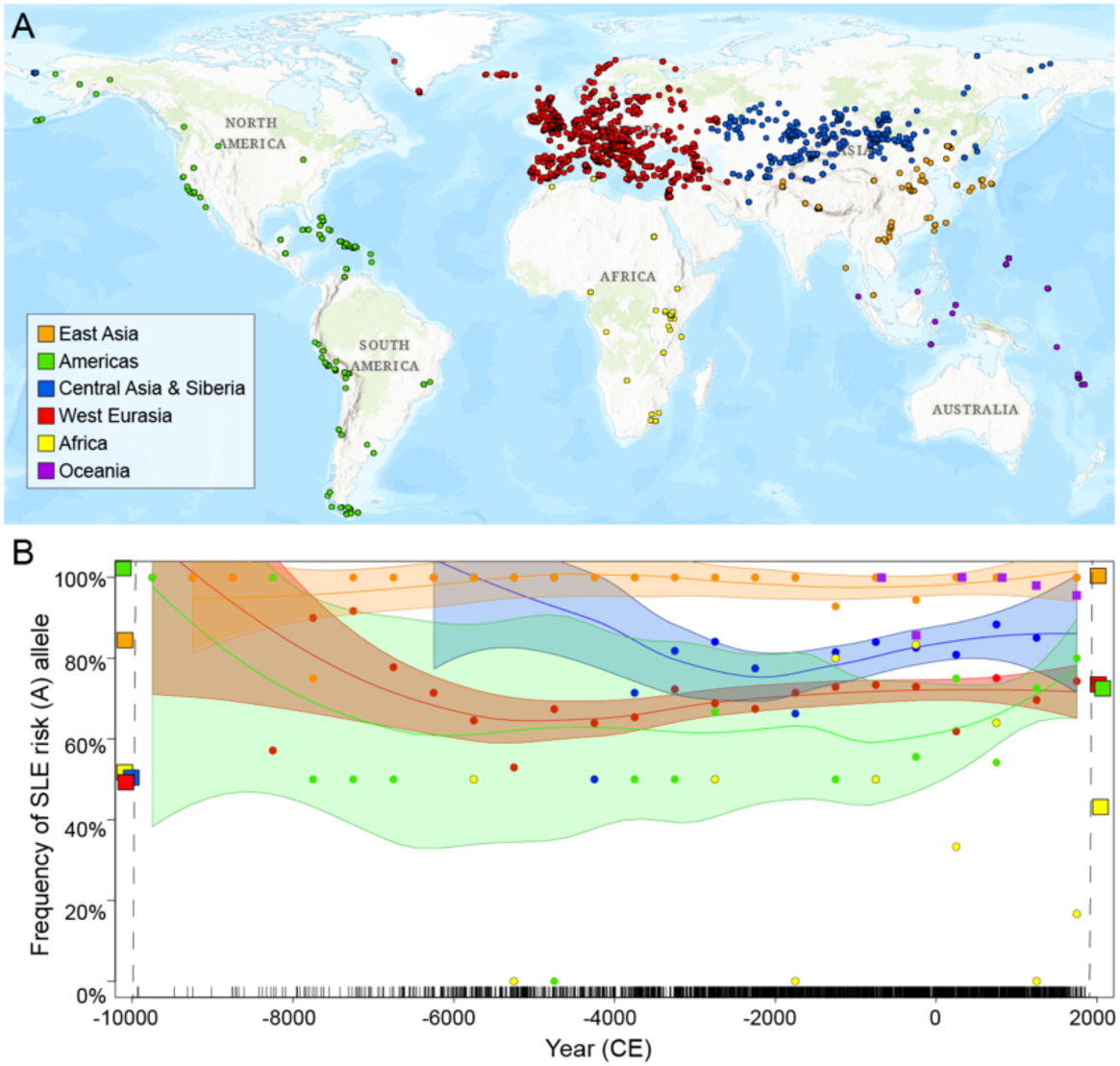
The *IRF7* lupus risk haplotype containing rs1131665 and rs1061502 had high population frequencies in the oldest human samples available, and these high frequencies are maintained over time across populations. (A) Global map of ancient DNA sampling locations included in this analysis (compiled from the Ancient Genome Diversity Project). Colors indicate ancestries used in analyses (see Methods). Sampling is most comprehensive in West Eurasia (Red) and Central Asia and Siberia (Blue) due to long-term preservation of DNA in colder climates. World map is depicted using a Cylindrical Equal Area Projection. (B) Plot of the mean population frequencies of the SLE risk haplotype for each 500-year bin are shown (dots), and each population is fit with a weighted loess curve and 95% confidence intervals to show trends of the frequency of the SLE risk allele over time. Confidence intervals for Oceania and Africa were not calculated due to small sample sizes. Genomes older than 10,000 BCE were sparse (N=36 across all ancestries) and were not included in the analysis. Instead, mean frequencies of the haplotype in each ancestry between 43,000 BCE and 10,000 BCE are represented by boxes on the left-hand y-axis. Current allele frequencies are shown as boxes on the right (taken from Ensembl). Each black vertical line along the x-axis represents an ancient DNA sample that passed quality control measures as described in the Methods section. Ancestry colors are the same as in panel A.

### The lupus-associated *IRF7* haplotype increases IFN-α responses downstream of TLR-7

To gain a biological understanding of the *IRF7* lupus risk haplotype, we next examined the effects of the haplotype on IRF7 activity. IRF7 is activated downstream of numerous PRR pathways^15,16,18,44^, resulting in nuclear localization, binding to DNA, and subsequent transcription of IFN-I **(Figure 3A).** We hypothesized that the lupus risk haplotype would increase interferon stimulated regulatory element (ISRE) activity and IFN-I production downstream of these innate immune pathways.

**Figure 3.**
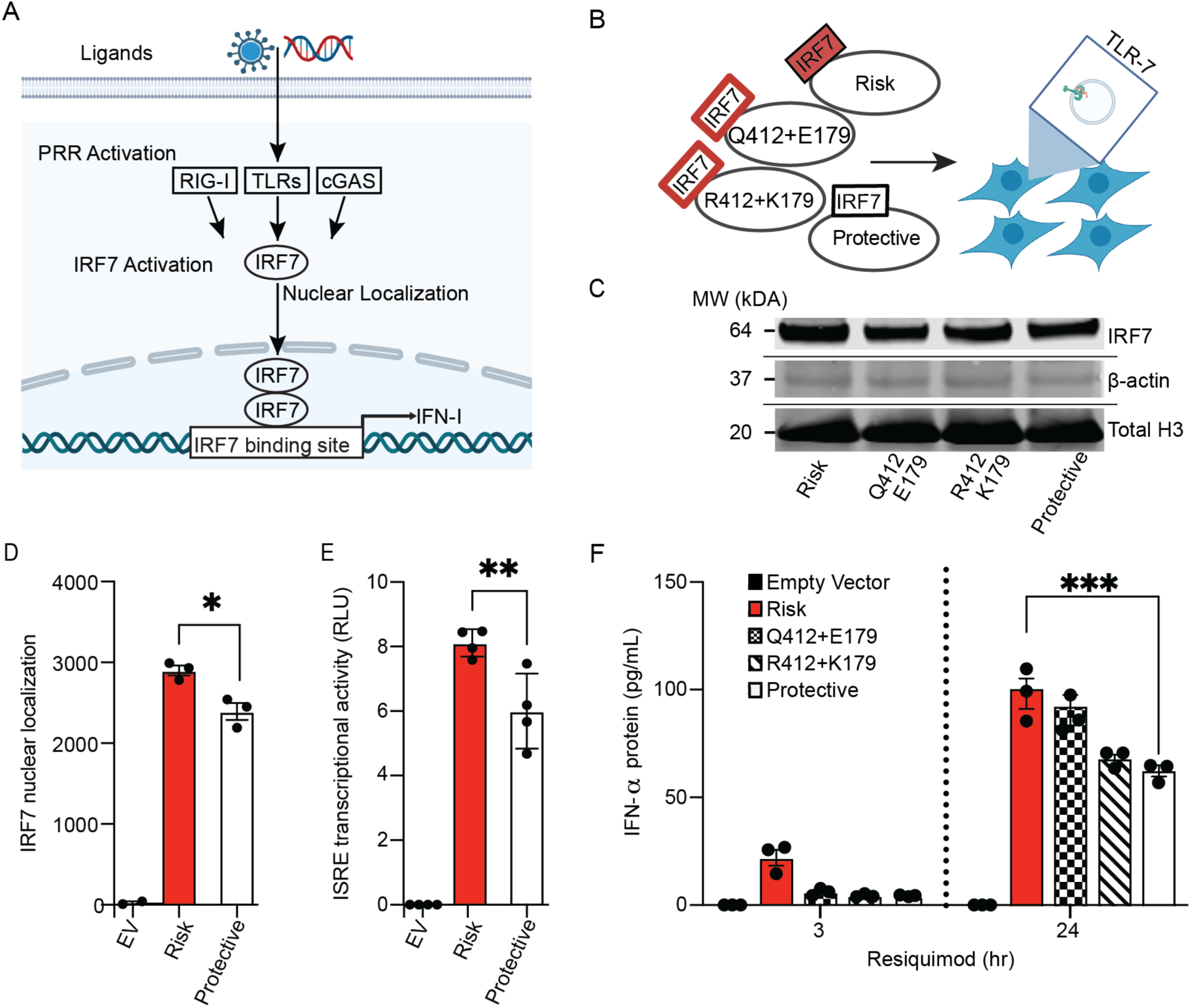
HEK Blue hTLR7 cells transfected with the *IRF7* lupus risk haplotype exhibit higher IFN-α induction and secretion compared to other allelic combinations. (A) Schematic of IRF7 activation, nuclear localization, and DNA binding downstream of multiple pattern recognition receptor pathways. (B) Overview of experimental approach employed in HEK Blue hTLR7 cells. C) Representative Western blot image of HEK Blue hTLR7 nuclear lysates following stimulation with R848 for 3-hours. β-actin and total H3 are included as loading controls. (D) Densitometry analysis of band intensities for risk and protective haplotype bands based on the quantitative Western blot presented in Figure S2. IRF7 bands were normalized to a total protein stain to calculate normalized signal as described in Methods. Welch’s t-test was used to determine significance. (E) Normalized ISRE luciferase activity (ratio of ISRE firefly to Renilla luciferase relative light units (RLU)) following 24-hours of R848 stimulation. Welch’s t-test was used to determine significance. (F) IFN-α measured in cell-free supernatants from cells stimulated with R848 for 3– and 24-hours. Data are mean ± SEM of triplicate samples (unless otherwise indicated) and are representative of at least three independent experiments. Significance was assessed with two-way ANOVA with a Holm-Šidak’s multiple comparisons correction. *P<0.05; ***P<0.005; ns indicates non-significant (P>0.05). ISRE: Interferon stimulated response element; EV: Empty Vector; R848: Resiquimod.

We first assessed IRF7 activity downstream of endosomal Toll-like Receptor-7 (TLR7), which is activated by host and viral nucleic acids ^45,46^. The role of TLR7 signaling in the etiology of lupus is well-established ^47^, and TLR7 gain-of-function and increased signaling alone is sufficient to cause a monogenic form of lupus ^48^. We employed a system using cells expressing one of the four possible combinations of IRF7 genotypes in HEK cells that stably express TLR7 (HEK Blue hTLR7) **(Figure 3B**). Since one of the amino acid changes within the risk haplotype (Q412R) is located within the IRF7 inhibitory domain **(Figure 1D)**, which controls IRF7 nuclear localization ^21^, we hypothesized that the risk haplotype would increase nuclear localization of IRF7 downstream of TLR7. Indeed, when we stimulated HEK Blue hTLR7 cells expressing the IRF7 constructs with TLR7/8 agonist Resiquimod (R848) and probed IRF7 in the nuclear lysates, we observed a similar level of total IRF7 protein (**Figure 3C; Figure S2)** and increased nuclear localization of IRF7 in the risk haplotype compared to the protective haplotype **(Figure 3D, Figure S2)**. To determine if this difference in nuclear localization leads to genotype-dependent transcriptional activity, we next assessed R848-stimulated cells expressing the risk or protective haplotype of IRF7 in the context of a luciferase reporter containing three copies of the interferon stimulated response element (ISRE) regulatory sequence recognized by IRF7. We observed genotype-dependent ISRE regulatory activity, with the risk haplotype leading to increased ISRE transcriptional activity compared to the protective haplotype **(Figure 3E)**. We next assessed whether this increased IRF7 activity leads to increased IFN-α protein secretion. Strikingly, we observed robust genotype-dependent differences in IFN-α production after stimulation with R848 **(Figure 3F)**.

When we assessed combinations of the two mixed haplotypes (that rarely occur in humans), we found that neither combination fully replicated the increase in IRF7 nuclear localization observed for the risk-only compared to the protective-only haplotype, with the largest change coming from the 412 risk (Q) variant **(Figure 3C, Figure S2).** We further found that the risk allele of the risk Q412 variant alone results in genotype-dependent transcription and translation of type I interferons, as well as a 2-fold increase in ISRE activity relative to R412 **(Figure S3 A-B).** Consistent with the IFN-I findings, we found that Q412R also leads to >2-fold increased IFN-I responsive gene expression (e.g., *OAS2, IFIT1, IFIT3,* and *CXCL10* transcript levels based on RNA-seq experiments in HEK Blue hTLR7 cells – see Methods) **(Figure S3C-G**).

Based on the additive biological impact of the two variants and the fact that the risk alleles of the haplotype are in perfect linkage disequilibrium and almost always co-occur in humans, we conclude that the risk haplotype contributes to heightened TLR7-dependent IFN-I production. Altogether, the results of these experiments establish that the lupus risk haplotype increases IRF7 nuclear localization, ISRE-based transcriptional regulation, and IFN-I secretion relative to the lupus protective haplotype.

### THP-1 monocytes stably expressing physiological levels of IRF7 exhibit genotype-dependent interferon production

Given our results in a transient transfection system in HEK Blue hTLR7 cells, we next assessed the effects of the *IRF7* lupus risk haplotype in monocytes, a cell type that is critical to the IFN-I signature in lupus ^49,50^. THP-1 monocytes were transduced at a multiplicity of infection (MOI) that results in IRF7 expression levels approximating physiological levels (**Figure 4A, Figure S4**). The IRF7 constructs used in this study employed an HA epitope tag and were tested for IFN-α induction downstream of TLR7 activation to ensure sustained IRF7 transcriptional activity **(Figure S5**). Previous studies have shown that THP-1 monocytes produce IRF7-dependent IFN-α in response to a variety of agonists, including Poly(I:C) ^51,52^. We confirmed that Poly(I:C) transfection leads to IRF7 phosphorylation in our system and that risk and protective monocytes express equal amounts of total IRF7 protein (**Figure S6**). THP-1 monocytes expressing the lupus risk haplotype produced >2-fold more IFN-α mRNA compared to the lupus protective haplotype at 3-hours (P<0.05) and 12-hours (P<0.01) post-transfection **(Figure 4B).** The genotype-dependence of increased IFN-α was attenuated and no longer statistically significant at 24 hours, likely due to post-transcriptional negative feedback of the signaling pathway **(Figure 4B).** The interferon-stimulated gene (ISG) response of THP-1 monocytes to exogenous universal IFN-I administration was not fully dependent on the presence of IRF7 protein **(Figure S7**), consistent with previous studies ^53^. This suggests that the IRF7 lupus risk haplotype amplifies the primary production of IFN-α, with indirect amplification of ISGs downstream of this IFN-I secretion. We also examined secreted IFN-α in our system. At 3 hours post transfection with Poly(I:C), no detectable IFN-α protein was observed **(Figure 4C)**, consistent with the relatively low levels of mRNA induction at this time point **(Figure 4B).** Twenty-four hours after transfection, we observed a >2-fold (P<0.0001) increase of IFN-α protein production in cells expressing the risk haplotype compared to the protective haplotype **(Figure 4C).** This two-fold difference is consistent with our data obtained in the HEK Blue hTLR7 system as well as previous data ^33^. As IRF7 deficient cells failed to produce detectable IFN-α, these results support the role of IRF7 as the central regulator of IFN-α production downstream of PRR pathways in this cellular context.

**Figure 4.**
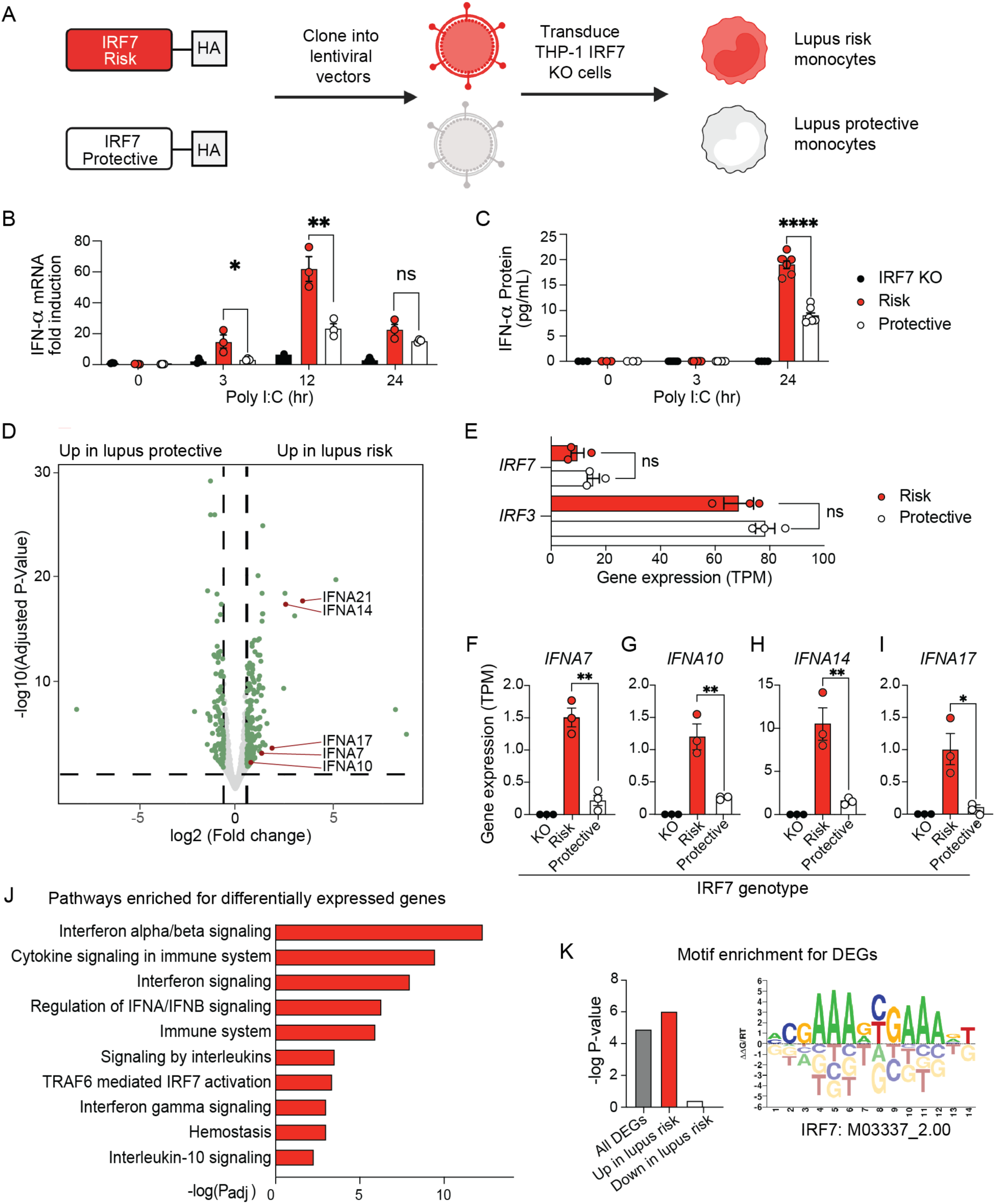
THP-1 monocytes stably expressing the *IRF7* lupus risk haplotype exhibit higher IFN-α induction at the transcript and protein levels. (A) Schematic showing the generation of THP-1 monocytes stably expressing the lupus risk and protective haplotypes in *IRF7*. (B) Quantitative PCR analysis of mRNA expression of IFN-α (subtypes IFNα2, IFNα25, and IFNα21) following transfection with Poly(I:C) at multiple time points in IRF7-deficient (KO) cells engineered to express the risk or protective haplotypes of *IRF7*. (C) IFN-α measured in cell-free supernatants from cells transfected with Poly(I:C). (D) Differentially expressed genes in THP-1 monocytes expressing either the lupus risk or the lupus protective haplotypes 3-hours post-transfection of Poly(I:C). (E) Gene expression profiles [transcripts per million (TPM) values] of *IRF3* and *IRF7* in lupus risk and lupus protective monocytes. (FI) Gene expression profiles [TPM values] of IFNA subset genes differentially expressed in IRF7 deficient (KO), lupus protective, and lupus risk monocytes 3-hours post transfection with Poly(I:C). (J) Pathway enrichment analysis of differentially expressed genes from D. (K) Motif enrichment for a representative IRF7 DNA binding motif for genes differentially expressed in the analysis presented in D. The motif image is taken from CIS-BP data build 2.00. Data are mean ± SEM of triplicate samples and are representative of at least three independent experiments, except the RNA-seq experiment in Figure 4D which was performed once for all replicates. Statistical comparisons between risk and protective are based on a two-tailed unpaired Student’s t-test; *P<0.05; **P<0.01; ****P<0.0001; ns indicates non-significant (P>0.05).

To assess transcriptional differences driven by the lupus haplotype in an unbiased manner, we next performed RNA-seq on lupus risk, lupus protective, and IRF7-deficient monocytes 3-hours post-transfection with Poly(I:C) **(Tables S2 and S3).** As expected, Poly(I:C) stimulation is a bigger source of variation than genotype in these experiments **(Figure S8).** We observed 306 differentially expressed genes (DEGs, alpha threshold α=0.05, 1.5-fold or greater difference in expression levels), with 200 genes upregulated in the lupus risk haplotype and 106 genes upregulated in the protective haplotype **(Figure 4D, Table S4)**. This genotype-dependence was not due to differences in *IRF7* or *IRF3* expression levels, as these genes exhibit similar expression levels in both genotypes of our experimental system **(Figure 4E).** We observed significant upregulation of multiple *IFNA* transcripts in the lupus risk haplotype compared to the protective haplotype **(Figure 4F-I)**, and IRF7 deficient cells did not produce *IFNA* transcripts, consistent with an IRF7 intrinsic effect. Pathway enrichment analysis of DEGs revealed significant enrichment for the “Interferon alpha/beta signaling” (P_adj_= 3.9×10^−13^), “Interferon signaling” (P_adj_= 8.7×10^−9^), and “IRF7 activation” (P_adj_=3.2×10^−4^) pathways (**Figure 4J, Table S5)**. When pathway analysis was performed separately on genes upregulated by the risk and protective haplotypes, we observed a similar pattern of enrichment in those genes upregulated by the risk haplotype, while genes upregulated by the protective haplotype were not enriched for these pathways **(Figure S9).** The promoters of DEGs upregulated by the risk haplotype were significantly enriched for predicted IRF7 binding sites, consistent with a mechanism involving stronger DNA binding of lupus risk IRF7 leading to stronger induction of gene expression **(Figure 4K)**. Taken together, these data suggest that the IRF7 lupus risk haplotype drives a distinct transcriptional program in monocytes characterized by roughly two-fold higher IFN-α RNA and protein levels than the protective haplotype.

### The *IRF7* lupus risk haplotype increases DNA binding strength and alters the DNA sequence specificity of IRF7

Next, we sought to identify allele-dependent differences in IRF7 DNA binding. To this end, we used the Protein Binding Microarray (PBM) technology ^54^. This array-based method measures the DNA binding of a tagged transcription factor to thousands of double stranded DNA probes, providing a comprehensive survey of DNA-binding strength and specificity for a given protein. We used a previously published PBM array design that contains thousands of DNA elements recognized by IRF proteins (see **Methods**) ^55^. Because IRF7 requires post-translational modifications, and because we observed allele-dependent IRF7 nuclear localization **(Figure 3C and D)**, we used the nextPBM technology, which profiles DNA binding using nuclear extracts ^56^. We performed nextPBM experiments using extracts from untreated or Poly(I:C) transfected THP-1 monocytes expressing the IRF7 lupus risk haplotype or the protective haplotype attached to a HA-tag, as employed in the experiments in **Figure 4**.

The PBM probe intensities for the IRF7 lupus risk haplotype after Poly(I:C) transfection correlate strongly with those obtained in a previously published study for purified IRF7 homodimers (Pearson correlation coefficient (PCC) = 0.95) ^55^ (**Figure 5A, Figure S10**). The motif derived from the risk haplotype in the transfected condition matches the known motif for the IRF7 homodimer (**Figure S11**). DNA binding of the IRF7 lupus risk haplotype in the untreated condition was weaker than the Poly(I:C) condition, in agreement with lower levels of nuclear IRF7 in this condition (**Figure S10**), and was still highly concordant with IRF7 homodimer binding (PCC = 0.89). Strikingly, the binding strength of the IRF7 lupus risk and protective haplotypes were nearly identical in untreated conditions (**Figure S10**). In contrast, IRF7 binding strength for these haplotypes deviated dramatically with Poly(I:C) transfection, with a significant increase in binding of the risk haplotype to the IFN-type probes (2bp-type ISRE elements) but little change for the protective haplotype (**Figure 5B, Figure S10).** Distinct DNA binding preferences for the IRF7 lupus risk and protective haplotypes were observed for elements with 2– or 3-bp spacers between 5’-GAAA core elements (i.e., 5’-GAAAXXGAAA-3’ versus 5’-GAAANNNGAAA-3’) (**Figure 5B**). Further, motif analysis confirmed that IRF7 lupus risk haplotype binding agrees with that of the IRF7 homodimer, while the protective haplotype exhibits distinct base preferences (**Figure S11**). In particular, for the IRF7 lupus risk haplotype and IRF7 homodimer, we found stronger preference for a “C” nucleotide at positions 5 and 11, and a weaker preference for a “G” at position 16. Collectively, these results indicate that, in the Poly(I:C) transfected condition, the lupus risk-associated IRF7 binds more strongly to IFN-type sites in a manner consistent with an IRF7 homodimer. In contrast, while the lupus protective IRF7 also accumulates in the nucleus upon Poly(I:C) transfection, it exhibits weaker binding to IFN-type sites than the risk haplotype, along with altered DNA preferences at several base positions. While this could reflect direct changes in IRF7 interaction with DNA, the amino acid distinctions between risk and protective IRF7 variants may also alter interactions with other proteins in the nuclear extracts that shape DNA binding characteristics.

**Figure 5.**
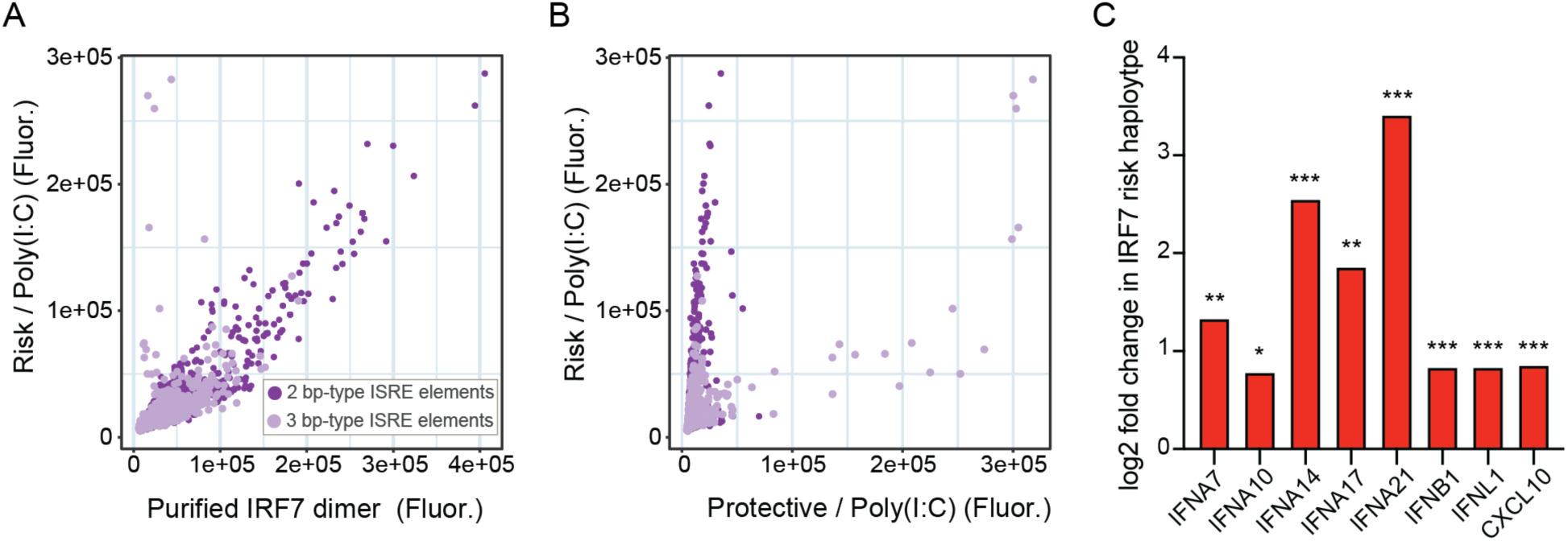
The *IRF7* lupus risk haplotype has increased DNA binding strength and altered DNA binding specificity compared to the protective haplotype. Comparison of DNA binding strength for the IRF7 lupus risk haplotype to purified IRF7 dimer (A) or the IRF7 protective haplotype (B). Binding for IRF7 lupus risk and protective haplotypes was determined via nextPBM assays using extracts from lupus risk and protective THP-1 monocytes following 3-hours of Poly(I:C) transfection. Data for the purified IRF7 dimer is from Andrilenas et al. 55, which used a mutant form of IRF7 that is constitutively dimeric. Binding is shown to 6,954 2-bp-spacer variant IRF7 binding sites (dark) and 3,456 3-bp-spacer variants (light) (see Methods) and is quantified by median PBM probe fluorescence values for each DNA sequence. (C) Increased binding of lupus risk haplotype IRF7 to the promoters of genes assessed on the PBM leads to increased gene expression (RNA-seq) of the 8 indicated genes. Log2 fold change values for these genes are plotted along with the adjusted P-value from the RNA-seq analysis (Figure 4). *Padj<0.005, **Padj<0.0005, ***Padj<1.0×10-5.

We next compared allele-dependent IRF7 DNA binding events to allele-dependent gene expression levels. To this end, we used the subset of PBM array probes that contain DNA sequences representing the promoters of 26 previously identified interferon-responsive genes ^55^. Among these 26 genes, 9 had significantly higher expression levels in the IRF7 lupus risk versus protective haplotype RNA-seq data, 4 were unchanged, none had lower expression, and 13 were not detected in THP-1 cells. These data are consistent with prior studies showing that THP-1 cells contain deletions in *IFNA* genomic regions, including *IFNA1*, *IFNA2*, *IFNA5*, *IFNA6*, *IFNA8*, and *IFNA13* ^57^. Strikingly, 8 of the 9 “high in lupus risk” genes also have significantly stronger IRF7 risk haplotype binding to their promoters in Poly(I:C) transfected conditions **(Figure 5C)**. Stronger binding of risk IRF7 to select interferon-responsive genes is also reflected at the protein level, with increased secretion of IFNA7, IFNA10, IFNA14, IFNA17, and IFNA21 manifested by the lupus risk compared to protective haplotype **(Figure 4D).** In contrast, only 1 of the 4 “unchanged” genes had higher IRF7 lupus risk haplotype binding (P=0.011, one sided proportions test). These results suggest that the stronger overall binding of the IRF7 lupus risk haplotype directly impacts gene expression levels, leading to elevated expression of IFN-I genes including *IFNA10* and *IFNB1*.

### Human lung epithelial cells expressing the lupus risk IRF7 haplotype exhibit genotype-dependent IFN-α secretion

The IFN-I response is critical to mounting effective immune responses, especially in response to viruses ^13^. We therefore hypothesized that the *IRF7* lupus risk haplotype could also increase IFN-I responses in airway epithelial cells ^25,58^. We generated A549 cell lines stably expressing the risk and protective haplotypes using the same lentiviral vectors employed in the THP-1 monocyte system, starting with A549 cells deficient in IRF7 **(Figure S12,** see **Methods)**. Because we could not use a chemical selection strategy, we performed single cell cloning to generate three independent clones of both the risk and protective haplotypes. Because A549 cells produce IRF7-dependent IFN-α in response to transfection of Poly(dA:dT) ^44,59,60^, we hypothesized that the risk haplotype would increase secreted IFN-α, as we observed with our monocyte and HEK Blue hTLR7 systems. Indeed, we again observed a 2-fold (P<0.005) difference in secreted IFN-α between the risk and protective haplotypes 24-hours after transfection with Poly(dA:dT) **(Figure S12B)**, with no detectable IFN-α produced in the IRF7 deficient cells.

### An engineered risk variant in mouse *Irf7* enhances antiviral immunity and autoantibody production in a lupus model

Human IRF7 has 69% amino acid identity with murine IRF7 ^61^. Importantly, the highest amino acid identity (>95%) is found in the DNA-binding and rs1131665-bearing inhibitory domains of IRF7. All inbred laboratory strains (e.g., C57BL/6J, BALB/cJ, 129S1/SvImJ, C3H/HeJ), as well as those genetically susceptible to autoimmunity (e.g., SJL/J and MRL/MpJ), are homozygous for the protective C/G allele at the mouse position orthologous to the human IRF7 variant rs1131665. Thus, both humans and mice with the protective variant are characterized by an arginine residue in the IRF7 inhibitory domain at positions 412 and 334, respectively. Using CRISPR targeting and homologous recombination, we mutated C/G to T/A at position 141,263,661 on chromosome 7 (Genome Reference Consortium Mouse Build 38 or mm10), resulting in an R334Q substitution in mouse IRF7 that is homologous to the rs1131665 SLE risk-variant R412Q substitution in human IRF7.

To test the hypothesis that homologous lupus-risk variants in mouse *Irf7* would exacerbate risk for autoimmune disease, we applied an epicutaneous resiquimod (R848) model that can trigger lupus-like autoimmunity in non-autoimmune prone mouse strains (e.g., C57BL/6) ^62,63^ (**Figure 6A**). After eight weeks of R848 treatment, mice with the lupus-risk IRF7 homologue (QQ) developed significantly more anti-double stranded DNA autoantibodies compared to mice with the lupus-protective IRF7 homologue (**Figure 6B**).

**Figure 6.**
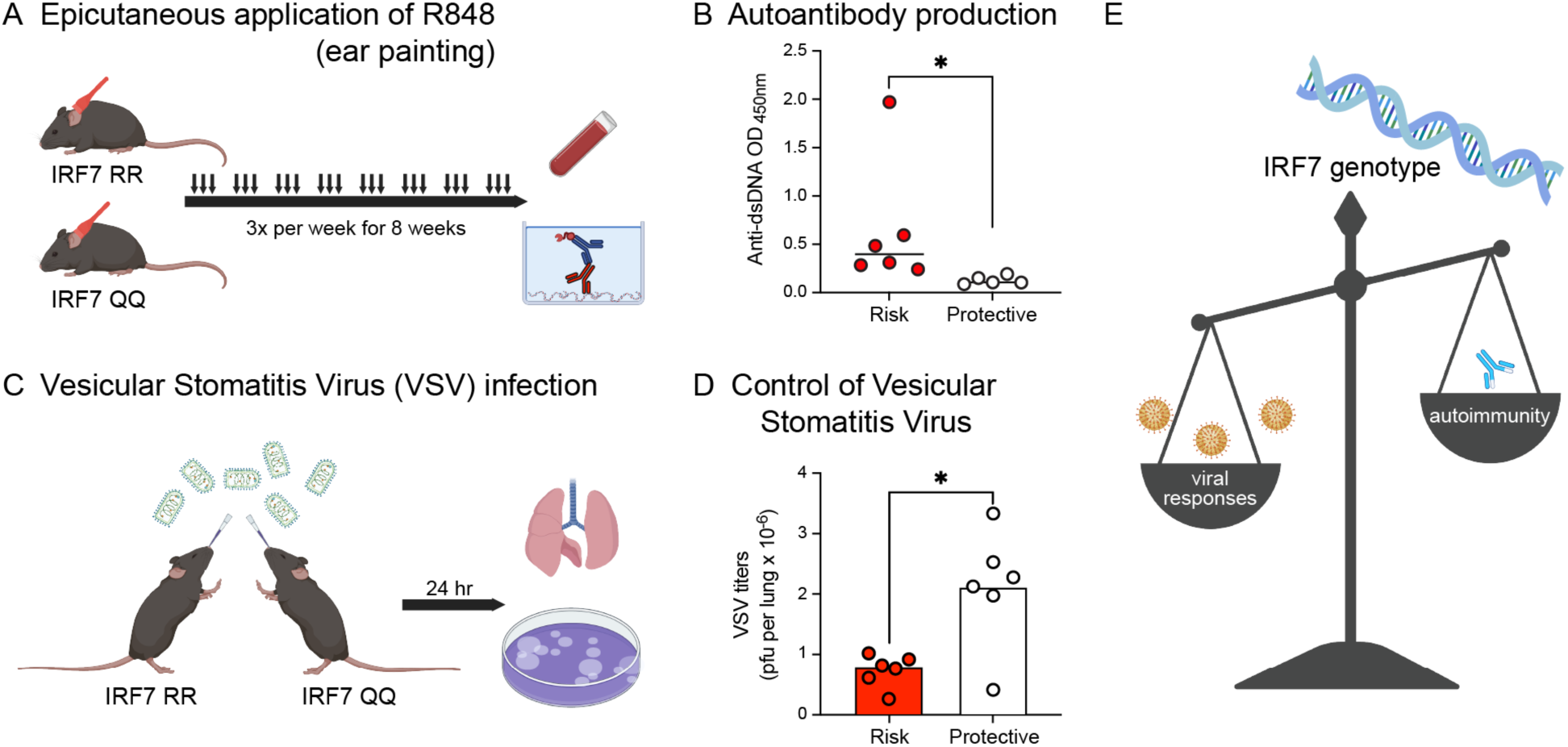
Mice with the *IRF7* lupus risk variant develop autoantibodies in a model of lupus and have a lower viral titer with Vesicular stomatitis virus infection. Using CRISPR targeting and homologous recombination, we mutated C/G to T/A at position 141,263,661 on chromosome 7 (Genome Reference Consortium Mouse Build 38), resulting in an R334Q substitution in mouse IRF7 that is analogous to the rs1131665 SLE risk-variant R412Q substitution in human IRF7. (A) 100 micrograms of resiquimod (R848) was epicutaneously applied three times per week to the ears of genome edited C57BL/6 mice with the lupus risk or non-risk IRF7 genotype for a total of eight weeks. (B) Sera was collected and used to quantify anti-double-stranded DNA (ds-DNA) antibodies by ELISA. (C) Eightweek-old male mice with the lupus risk or non-risk IRF7 genotype were intranasally infected with 1 million plaque-forming units of VSV-NJ. (D) After 24 hours, virus was quantified in lung homogenates using viral plaque assays. (E) Model for the lupus risk genotype in IRF7 enhancing both viral responses (with the lupus risk genotype enhancing viral clearance) and autoimmunity (with the lupus risk genotype enhancing the production of autoantibodies).

We next tested the hypothesis that the *Irf7* lupus-risk homologue could enhance innate immunity against virus infection (**Figure 6C**). After pulmonary infections with the interferon-sensitive vesicular stomatitis virus (VSV), IFN-sufficient mice (e.g., C57BL/6) rapidly eliminate replicating virus from the lungs in an IFN-dependent manner ^15,64^. We found that mice with the lupus-risk IRF7 homologue (QQ) exhibited significantly lower titers of VSV in lungs one day after infection compared to non-risk RR control mice (**Figure 6D**).

Taken together, these results suggest that the lupus risk haplotype increases IRF7 activity in a manner that can enhance antiviral defense but also increase risk for autoimmunity (**Figure 6E**). These results offer a plausible biological explanation for the strong conservation of this haplotype in ancient DNA **(Figure 2)**.

## DISCUSSION

Lupus arises from a convergence of genetic and environmental factors. The mechanisms by which specific risk alleles perturb immune regulation remain incompletely defined ^65–67^. Here, we focused on the highly prevalent *IRF7* coding haplotype associated with lupus risk in modern populations and conserved in ancient genomes, delineating its impact on IRF7-dependent IFN-I responses. Across multiple cellular contexts, the lupus risk haplotype increases IRF7 gene regulatory activity, culminating in augmented IFN-α transcription and secretion.

There is substantial evidence that IFN-I drives disease in lupus and plays a central role in the control of many viral infections ^49,50,68^. Consistent with this dual role, mice harboring the IRF7 lupus risk variant that enhances IFN production exhibited both elevated autoantibody titers in response to autoimmune triggers and improved control of pulmonary virus infections. Our data are therefore consistent with a model in which this highly prevalent haplotype exerts context-dependent effects, increasing risk for lupus and autoimmunity while simultaneously improving antiviral defense. This underscores the complex nature of coding genetic variants in the setting of immunoregulation and autoimmune disease, where variants that are harmful in one context may be beneficial in another.

IRF7 itself is an interferon-responsive gene whose expression is upregulated downstream of IFN-I signaling. In this study, we introduced variants of IRF7 whose expression was not altered by IFN-I so that we could decouple the immunologic consequences of increased IRF7 transcription factor activity from those of increased IFN-I production and signaling. Using this system, we observed consistent ∼2-fold or greater amplification of IFN-I production from cells expressing the lupus risk haplotype compared to those with the protective haplotype. In physiological systems, where increased IFN-I would be expected to further induce IRF7 expression, these primary differences are likely amplified through positive feedback loops that increase IRF7 levels and enhance IFN responses.

In models of autoimmunity and in patients with lupus, IRF7 is involved in the enhanced production of IFN-I, contributing to autoantibody production, tissue damage, and inflammation ^31,33,34,69^. Our findings extend these observations by demonstrating that a common *IRF7* coding haplotype quantitatively tunes IFN-I output in cell types central to lupus pathogenesis, including monocytes and airway epithelial cells, and that the corresponding murine homologue augments autoantibody production in a TLR-7–driven lupus model. These results fit within a broader framework in which endosomal TLR7 gain-of-function is sufficient to cause lupus in humans ^48^, and in which monocytes and plasmacytoid dendritic cells act as major sources of IRF7-dependent IFN-I in both infection and autoimmunity ^49,50,68^. Amplified IFN-I can arise downstream of both pathogens and self-nucleic acids, each contributing to chronic pathway activation in lupus ^70–72^. IRF7-mediated type I interferon signaling promotes lupus across compartments, driving autoreactive germinal center and plasma cell responses in B cells ^73^. In parallel, IRF7 and STAT1 are required for control of infection with IFN-sensitive viruses, including VSV and influenza A virus ^52,74^, consistent with our observation that the lupus risk Irf7 allele increases innate control of pulmonary VSV infection.

IRF7 thus emerges as a key nodal point integrating genetic risk for lupus with host defense against viral pathogens. Loss-of-function mutations in IRF7 affect susceptibility to viral infections such as SARS-CoV-2, influenza A virus, and hepatitis C by attenuating transcriptional amplification of type I IFN genes ^24,26,27^. At the opposite end of this functional spectrum, our data indicate that a gain-of-function IRF7 haplotype enhances IFN-I induction and accelerates viral clearance, but at the cost of increased autoantibody production. We initially hypothesized that a selective pressure may have led to the increased *IRF7* lupus risk frequency in the East Asian ancestry dataset (98%) compared to African ancestry (42%). However, the risk allele frequency was consistently high throughout the sampling period starting from the Antonine Plague of CE 165 to 180 in the Roman Empire and including the cycles of Bubonic Plague in the 14^th^-18^th^ centuries. Thus, the change in frequency from a presumably lower ancestral population must have occurred before the time frame captured in ancient DNA genomes. The protective role of the lupus risk haplotype of IRF7 that we observe in IFN-I production is consistent with both the effect size of increased lupus risk and the persistence of this haplotype in human populations over time.

## RESOURCE AVAILABILITY

Lead Contact: Leah Kottyan.

Materials Availability: Cell lines and constructs generated in this study are available upon request. Data and Code Availability: All RNA-seq data will be made available upon publication.

## Data Availability

All data produced in the present study are available upon reasonable request to the authors

## ACKNOWLEDGEMENTS

The authors would like to acknowledge the CCHMC Genomics Sequencing Facility (RRID:SCR_022630) and the CCHMC Viral Vector Shared Facility (RRID: SCR_022641). This research was funded by National Institutes of Health (NIH) R01 AR073228 (to L.C.K, M.T.W, and S.N.W), R01 DK107502, R01 AI148276, U01 HG011172, and U19 AI070235 to L.C.K.; R01 HG010730, R01 GM055479, U01 AI130830, U01 AI150748, P01 AI150585, and U24 HG013078 to M.T.W.; R01 AI176519 and Department of Defense Impact Award to S.N.W.; and P30 AR070549, R01 NS099068, and R01 AI024717 to M.T.W. and L.C.K. U01 AI150748 supported ERM, BRR, MTW, and LCK. The Hopkins Lupus Cohort is supported by NIH Grant R01 DK134625. The Korean cohort was supported through the Basic Science Research Program through the National Research Foundation of Korea (NRF) funded by the Ministry of Education (NRF-2021R1A6A1A03038899). Funding was also supplied by the Office of Academic Affairs and Career Development to some trainees. VSV progenitor was a gift from David Masopust. Schematic figures were made in BioRender.

## AUTHOR CONTRIBUTIONS

S.J.V., S.N.W., M.T.W., and L.C.K. designed the study with contributions for N.S.. S.J.V. performed a majority of the experiments with experimental support from K.A.D., A.L., L.C, L.P., H.K.H., C.Y., O.G., A.A.D., S.J., C.F., M.H., Y.K., Y.B., B.R, and O.D. M.D. performed the PBM experiments. D.K. and B.A. performed in vivo VSV infection experiments. S.J.V., D.K., and B.A. performed mouse autoimmunity model experiments with support from H.S. S.J.V. and L.C.K. performed the fine mapping analysis with genetic data provided by the LLAS2 authors (H.C.A, M.E.C, S.B.G, A.A, S.K.N, A.M.S, B.IF., B.P.T, C.O.J, D.L.K, E.E.B, G.S.G, G.S.A., J.D.R, J.M.A, J.A.J, K.LS, L.A.C, L.M.V., M.E.A.R, M.P., R.H.S, R.P.K, R.R.G, S.C.B, S.A.B, D.C.G., T.J.V, J.M.G, C..D.L, P.M.G, J.A.K., G.D.G, K.M.K, and J.B.H). L.P.L. performed the evolutionary analysis. S.J.V. performed the RNA-seq analyses with analytical support from A.N.R, E.R.M, S.P., A.V., L.E.E., X.C. and M.T.W. T.S. supervised the PBM experiments and performed analysis of this data with M.T.W. S.J.V, L.P.L, L.C.K. and M.T.W. generated figures, performed data analysis, and wrote the manuscript with critical feedback from all authors. L.C.K., M.T.W., and S.N.W. provided funding and supervised the study. All authors approved the manuscript.

## SUPPLEMENTAL FIGURE TITLES AND LEGENDS

**Figure S1. rs1131665 (Q412R) and rs1061502 (K179E) account for all of the lupus risk (P<0.01) at the *IRF7-PHRF1* locus, related to Figure 1**. Each variant is represented as a data point in the context of its genomic location within the *IRF7-PHRF1* locus. Variants are colored based on their P-value, with red dots representing significant genetic variants (P<5×10-8) (none) and blue dots signifying variants that do not meet this threshold. (A-B) Step-wise logistical regression in the EU ancestry shows that the genotypes of these variants account for virtually all of the lupus association (P<0.01) at this locus. Admixture estimates were included as covariates in the logistic regression model.

**Figure S2. Unedited quantitative Western blot images of transiently transfected IRF7 haplotypes after 3-hours of stimulation with R848 in nuclear lysates of HEK Blue hTLR7 cells, related to Figure 3**. Each lane is denoted by the replicate number and the specific *IRF7* haplotype transfected. (A) IRF7 image following incubation with IRF7 antibody. (B) Total protein stain image stained with Revert 700 total protein stain (LI-COR biosciences). For densitometric analysis, each band for each replicate in image A was normalized to the band for each respective replicate in the total protein stain in image B to obtain normalized signal, as implemented in Empiria Studio software (LI-COR biosciences, see Methods). EV=Empty Vector, QK=Q412+K179, lupus risk haplotype; QE=Q412+E179; RK= R412+K179; RE=R412+E179, lupus protective haplotype.

**Figure S3. Transient transfection with IRF7 rs1131665 (Q412R) results in genotype-dependent IFN-I production at both mRNA and protein levels following TLR-7 stimulation with R848 in HEK Blue hTLR7 cells, related to Figure 3**. (A) IFN-α ELISA on cell-free supernatants collected at 3– and 24-hours post stimulation with R848 from cells transfected with Q412 or R412 *IRF7* variants. (B) ISRE activity (ratio of ISRE firefly to Renilla luciferase relative light units (RLU)) after 24-hours of stimulation with R848 in cells transiently transfected with Q412 and R412. (C) Differentially expressed genes in HEK Blue hTLR7 cells expressing either the lupus risk or the lupus protective haplotypes. (D-G) Gene expression profiles [TPM values from RNA-seq] of relevant ISG transcripts 3-hours post R848 stimulation in cells transiently transfected with Q412 or R412 *IRF7*. Statistics for B and C are based on an unpaired t-test. *P<0.05; **P<0.01; ***P<0.005. Statistics for D-G above each plot show p-values adjusted using the Benjamini–Hochberg procedure to control the false discovery rate at 5% in DESeq2.

**Figure S4. THP-1 cells expressing the lupus risk and protective IRF7 haplotypes express relevant levels of IRF7, related to Figure 4**. Quantitative PCR analysis of *IRF7* RNA from unstimulated risk and protective monocytes. Fold induction is calculated relative to the unstimulated endogenous THP-1 control. Data are mean ± SEM of duplicate samples.

**Figure S5. IRF7-HAx3 constructs do not disrupt IRF7 regulatory activity, related to Figure 4**. IFN-α ELISA on cell-free supernatants collected at 3-hours post stimulation with R848 from HEK Blue hTLR8 cells transfected with the risk IRF7 (Q412) tagged with eGFP on the N-terminal region, the C-terminal region, HAx3 on the C-terminal region, a construct containing IRF7 with no tag, or an untransfected control. Data are mean ± SEM of triplicate samples. GFP=Green fluorescent protein.

**Figure S6. Western blot analysis of IRF7 levels in whole cell lysate and nuclear pIRF7 in IRF7 lupus risk, protective, and IRF7 deficient monocytes, related to Figure 4**. Top: Total IRF7 in unstimulated whole cell lysates. β-actin is shown as a loading control. Bottom: pIRF7 in nuclear lysates 3-hours post-transfection with Poly(I:C) of THP-1 cells deficient in IRF7 (KO), expressing the lupus risk haplotype, or expressing the protective haplotype. Histone H3 is displayed as a loading control.

**Figure S7. IRF7 does not drive the response to IFN-I, related to Figure 4**. (A-B) Quantitative PCR of the IFN-I responsive genes (A) *OAS1* and (B) *MX1* following exogenous administration of 1000 units/mL of Universal IFN-I in KO (white) and IRF7-sufficient (red) THP-1 monocytes at 1-hour, 6-hours, and 18-hours post stimulation. Fold induction is calculated relative to the unstimulated control. Data are mean ± SEM of duplicate or triplicate samples.

**Figure S8. PCA of the 500 most variable genes in THP-1 RNA-seq experiments, related to Figure 4**. Principal Component Analysis (PCA) on the THP-1 RNA-seq datasets (Figure 4) used in this study. Samples were paired-end sequenced using a whole transcriptome library.

**Figure S9. Pathway enrichment analysis of genes upregulated by the IRF7 lupus risk haplotype and protective haplotypes at 3-hours post-transfection with Poly(I:C), related to Figure 4**. (A) Pathway analysis of genes upregulated by the lupus risk haplotype. (B) Pathway analysis of genes upregulated by the lupus protective haplotype.

**Figure S10. DNA binding of lupus risk and protective IRF7 compared to purified IRF7 homodimers at 3-hours post-transfection with Poly(I:C), related to Figure 5**. (A-D) Comparison of DNA binding strength for purified IRF7 dimer to the lupus risk (A-B) or protective (C-D) IRF7 from untreated (UT) (A,C) or Poly(I:C) transfected (B,D) THP-1 cell extracts. Binding strength is shown for 6,954 2-bp-spacer ISRE variant sites (see Methods). Binding is quantified by median PBM probe fluorescence values for each DNA sequence. Data for the purified IRF7 dimer are from Andrilenas et al., which used an IRF7 (8D) mutant that is constitutively dimeric. Insets show the same plots rescaled for direct comparison.

**Figure S11. DNA binding specificity differences of lupus risk and protective IRF7 at 3-hours post-transfection with Poly(I:C), related to Figure 5**. (A) PBM-derived motifs for purified IRF7 dimer, IRF7 lupus risk, and IRF7 protective proteins. Motifs are constructed by quantifying binding to a seed sequence (5’-TAACC<u>GAAA</u>CC<u>GAAA</u>CC TAA-3’, GAAA core elements underlined) and all single-nucleotide variants. (B) DNA binding of IRF7 lupus risk and IRF7 protective from Poly(I:C) transfected extracts to 10,410 IFN-derived DNA sequences (see Methods). Highlighted are the seed (red) and single-nucleotide variant (green) sequences used to define the binding motifs shown in (A).

**Figure S12. Clones of A549 cells expressing the *IRF7* lupus risk and lupus protective haplotypes demonstrate genotype-dependent IFN-α production, related to Figure 6**. A) Schematic showing the generation of A549 cells stably expressing the lupus IRF7 risk and protective haplotypes. B) IFN-α ELISA on cell-free supernatants collected at 24-hours post-transfection of A549 cells transfected with Poly(dA:dT). Bars are mean ±SEM of triplicate samples (Clones 2 and 3) or 6 samples (Clone 1). Significance of comparison between risk and protective haplotypes assessed through Linear Mixed-Effects Model with clone treated as a strata variable (P = 0.0162).

## STAR METHODS

### Genotyping of Genetic Variants

In this study, we genotyped 120 SNPs covering the *IRF7* region, spanning GRCh37 chr11: 550,028-697,661 as part of a larger collaborative study, the Large Lupus Association Study 2 (LLAS2). The samples were collected from individuals in the United States, Asia, Europe, and Latin America. They were genotyped using the Illumina iSelect platform located at the Lupus Genetics Studies Unit at the Oklahoma Medical Research Foundation. The subjects were grouped into four ancestral groups. All lupus patients met the American College of Rheumatology criteria for the classification of lupus ^75^ and were enrolled in this study through an informed consent process approved through the local Institutional Regulatory Boards. LLAS2 included genotyping of other lupus risk loci, and the analyses of those loci from this same collection, with and without lupus, have been published separately. Genetic variants were filtered to include only those with a call rate >0.95 and a Hardy-Weinberg P-value of <0.0001 in controls. Following variant filtration, samples that did not meet the >0.95 threshold were removed.

### Ascertainment of Population Stratification

The ancestries of the subjects in this study were self-identified. Genetic outliers from each ethnic and/or racial group were removed from further analysis as determined by principal component (PC) analysis and admixture estimates, as described previously ^76–78^. A PC analysis of the remaining samples (after outlier removal) confirmed that no sample had a PC1–3 more than 2 standard deviations outside of the mean. We used 347 ancestral informative markers (AIMs) from the same custom genotyping study that passed quality control in both EIGENSTRAT ^78^ and ADMIXMAP ^79,80^ to distinguish the four continental ancestral populations: Africans, Europeans, American Indians, and East Asians, allowing identification of the substructure within the sample set ^81,82^. We utilized PCs from EIGENSTRAT outputs to identify outliers of each of the first three PCs for the individual population clusters through visual inspection [see Fig. 1 of reference ^76^]. Three PCs were used because they accounted for 95% of the eigenvalues. Because the four admixture estimate proportions sum to 1, any three of the four provide the full set of information, so only three proportions were necessary to include as covariates; the EA proportion was omitted from this analysis, as it corresponded to the largest ethnic segment of the combined population. Admixture estimates were included as covariates in the stepwise logistic regression analysis.

### Genotype Imputation

To detect associated variants that were not directly genotyped, we imputed the *IRF7* region with Beagle 4.1 ^83^ using a composite imputation reference panel based on the 1,000 Genomes Project Phase 3 ^84^ sequence data. Imputed genotypes were included in the analysis if they met or exceeded a probability threshold of 0.5 and the same quality-control criteria thresholds described for the genotyped markers. The most likely genotype was used for variants passing quality controls in all analyses.

### Genetic Association Analysis

To identify groups with sufficient statistical power to identify true genetic associations, we performed a Power Calculation (α=0.05) for the Q412R haplotype. We did not assess genetic association at the *IRF7* locus in the East Asia and Americas ancestries due to insufficient power, as presented in Table S1.

We tested each variant for its association with lupus using logistic regression that included three admixture proportion estimates as covariates, as implemented in Golden Helix SVS (8.9.1). The additive genetic model was assessed as the initially tested model of inheritance. Using Golden Helix SVS, stepwise logistic regression was performed to identify those genetic variants independently associated with the development of lupus. For these analyses, admixture estimates were also used as covariates.

A trans-ancestral meta-analysis was performed using Golden Helix SVS (8.9.1). The P-values and odds ratios for each variant were included, and the analysis was weighted by the number of individuals with data for each variant in an individual cohort as part of an inverse variance model. For the trans-ancestral meta-analysis, only the European and African ancestral cohorts were used. Plots for genetic data were generated using locuszoomr (version 0.2.0) ^85^ and annotated using ensembldb (version 2.99.0) ^86^.

### Ancient DNA Analysis

A dataset containing ancient and present-day DNA was downloaded from the Ancient Genome Diversity Project ^87^ for haplotype analysis (16,389 unique individuals: 9,990 ancient, 6,399 present-day). This dataset contains ∼1.23 million positions in the genome (in hg19 coordinates) and is in a modified Eigensoft format. To extract haplotypes, the dataset was converted with the convertf program (EIGENSOFT 8.0.0 package; ^78,88^) into a ped file and then converted with PLINK^89^ into a BED file. All samples younger than 1,950 CE (age “0” years before present, reflecting radiocarbon date correction) were removed to only represent “ancient” samples. All ages were adjusted to “calendar year” for figures.

As this is not a whole genome dataset, but instead a sampling of SNPs across the genome, we searched for any lupus SNPs in perfect LD with the lead SNPs contained within this dataset. Of the SNPs within the lupus *IRF7* risk haplotype block, only the rs11246213 locus was present. This locus was extracted from the larger dataset, and then all samples with missing data at this locus were removed (ancient genomes have high proportions of missing data due to degradation compared to modern genomes). The dataset was also filtered to remove duplicates when a genome was sequenced more than once (common for ancient datasets as sequencing technologies improve). Almost all duplicates had the same rs11246213 haplotype across replicates, but a small subset had different alleles inferred (N=7). For these seven samples, we assessed data quality to choose the most probable haplotype and deleted the duplicate(s) (e.g., highest coverage from sequencing, or removing any replicates that indicated contamination).

The final dataset contained 4,503 individuals across Africa, Americas, East Asia, West Eurasia (European), Oceania, and Central Asia/Siberia. Most of the samples (particularly the very old samples) are from West Eurasia (Europe) and Central Asia/Siberia, because those are the places with permafrost and cold/deep caves to preserve very old skeletal remains including Neanderthal bones. Most samples are younger than 10,000 BCE, but the oldest are over 40,000 BCE in our final dataset. The haplotype for each individual was converted to an allele frequency percentage (0 for non-risk G, 0.5 for heterozygous, 1 for homozygous risk allele A). This dataset was then used to assess population allele frequencies from each ancestry in 500-year time bins. This bin size is appropriate based on the uncertainty inherent in radiocarbon dating due to the need to calibrate for background C-14 levels ^90^. Weighted loss curves were fit for each ancestry background from 10,000 BCE onward.

Human groups were assigned to ancestries based on definitions from the 1000 Genomes Project. The only populations that required additional attention were in the transition zone across Eurasia to divide populations into the historically relevant groups of (1) West Eurasia, (2) Central Asia and Siberia, (3) East Asia, and (4) Oceania. Recent ancient and modern DNA studies were used to assign populations across this region ^91–97^. The distribution of samples used in this study is depicted in **Figure 2A**.

### Cell Lines and Cell Culture

THP-1 IRF7 deficient and HEK Blue hTLR7 cells were purchased from InvivoGen. A549 IRF7 deficient cells were purchased from Abcam. THP-1 cells were grown in RPMI 1640 medium supplemented with 10% FBS, 10,000 units/mL of penicillin, 10,000 ug/mL of streptomycin, and Normocin. Following transduction (as described below), THP-1 cell lines stably expressing IRF7 variants were also cultured in 1 ug/mL puromycin. HEK Blue hTLR7 cells were grown in DMEM medium supplemented with 10% FBS, 10,000 units/mL of penicillin, and 10,000 ug/mL of streptomycin. A549 cells were grown in Ham’s F12-K (Kaighn’s) medium supplemented with 10% FBS, 10,000 units/mL of penicillin, and 10,000 ug/mL of streptomycin. All cells were kept at 37°C and 5% CO_2_ and were determined to be mycoplasma free.

### Expression Plasmid Generation

For stable IRF7 stable cell line generation, a pUNO1-hIRF7a-HA3x expression vector was purchased from InvivoGen and the region containing IRF7-HAx3 was cloned into a lentiviral transfer plasmid (pLentiPuro). The GeneArt site-directed mutagenesis system (Thermo Fisher Scientific) was used to generate the IRF7 lupus protective haplotype according to the manufacturer’s instructions. For transient transfections, an *IRF7* open reading frame was cloned into an overexpression vector from the Wolfgang Hammerschmidt lab, and the GeneArt site-directed mutagenesis system (Thermo Fisher Scientific) was also used to generate the various IRF7 mutants for this study according to the manufacturer’s instructions. All plasmids used in this study were verified using Sanger sequencing.

### Cell Stimulations and Treatments

Ligands used in cell stimulations include Resiquimod (R848) (Invivogen: 1ug/mL), polydA:dT-Lyovec (Invivogen: 2.5ug/mL), and Poly(I:C) LMW (Invivogen: 2.5ug/mL). Poly(I:C) was transfected using Lipofectamine 2000 (Invitrogen: 1ul/mL,). IFN-I stimulations used Human Universal Type I Interferon (PBL Assay Science: 1000 units/mL).

### Isolation of Nuclear Lysates for Western Blot

To obtain nuclear lysates for Western blots, respective cell lines were resuspended in 1 mL of cold PBS per 10 million cells. Cells were then centrifuged at 4°C, 300×g for 5 min and the PBS was aspirated off. Cell pellets were resuspended in 400μl of CE buffer (10 mM HEPES, pH 8; 10 mM KCl; 0.1 mM EDTA; 1 mM DTT; 1×Halt protease and phosphatase inhibitor) and incubated for 15 min on ice. Afterwards, 25μl of 10% Nonidet P-40 were mixed into the solution and the samples were centrifuged at max speed (17.3×g) at 4°C for 3 min. After discarding the supernatant, the cell pellet was resuspended in 30μl of NE Buffer (20 mM HEPES, pH 8; 0.4 M NaCl; 1 mM EDTA; 1 mM DTT; 1×Halt phosphatase inhibitor). Cells were sonicated using the Q125 sonicator (Qsonica) at 20% power, 5 s pulses for a total of 15 s. The supernatant was stored at –70°C. Protein concentration was measured using the bicinchoninic acid assay (BCA) (ThermoFisher Scientific) using a 5.5μl aliquot.

### Quantitative Western Blot Using Nuclear Lysates and Total Protein Stain

Nuclear lysates of samples were mixed with loading buffer and DTT and heated at 95°C for 2 min. After cooling, the samples were loaded into a 4–12% Nu page Bis–Tris Gel (Invitrogen) and ran for 90 min at 130 V in MOPS buffer, or 65 min at 130v in MES buffer. The gel was then transferred to a nitrocellulose membrane using the iBlot Machine (ThermoFisher Scientific). For Western blot normalization, membranes were stained using the Revert™ 700 Total Protein Stain (Licor Bioscience). The membrane was then blocked in Intercept Blocking Buffer for 1 h at room temperature. The primary antibody (IRF7: Santa-Cruz Biosciences) was diluted in Intercept Blocking Buffer (with Tween 20 diluted at 1:1000) and the membranes incubated with rocking in primary antibody overnight at 4°C. Membranes were washed in a PBS/Tween 20 solution twice and were incubated in fluorescently labeled secondary antibody in Intercept Blocking Buffer (with Tween 20 and SDS) for 60 min at room temperature with rocking. Membranes were washed again in a PBS/Tween 20 solution 2 times before being imaged using the Odyssey DLx Imaging System (Licor Bioscience). Protein expression levels were determined using Empiria Studio v2.2 (Licor Bioscience).

### Quantification of ISRE Transcriptional Activity Using Renilla Luciferase

IRF7 ORFs with the various amino acid changes were integrated as described above into an overexpression vector that was a gift from the Wolfgang Hammerschmidt lab. These plasmids expressing IRF7 were co-transfected (50 ng per well) into HEK Blue hTLR7 cells seeded in 96-well plates (30,000 cells per well) using TransIT-2020 Transfection Reagent (Mirus) with ISRE reporter (50ng per well) or a negative control plasmid (50 ng per well) without an ISRE reporter from BPS Bioscience according to the manufacturer’s protocol. Post-transfection, cells were stimulated for 24-hours with R848 and the two-step luciferase (Firefly & Renilla, BPS Bioscience) was used to quantify firefly and renilla luciferase signals. The firefly luciferase signal was normalized to the renilla signal for each replicate and then further normalized to the ISRE negative control signal to calculate normalized ISRE luciferase activity for each replicate.

### Quantification of Secreted IFNα

Quantification of secreted human IFNα from cell supernatants was measured by Human IFN-Alpha All Subtype ELISA Kit, High Sensitivity (Serum, Plasma, TCM) from PBL Assay Science.

### Lentiviral Vector Generation

Lentiviral transfer plasmids (pLentiPuro) containing IRF7 ORFs were given to the Viral Vector Core at Cincinnati Children’s Hospital for generation of VSV-G pseudotyped lentiviral vector generation. Viral titer was determined by puromycin titration.

### Transduction of IRF7 Into Monocyte and Airway Epithelial Cell Lines

THP-1 IRF7 deficient cells were transduced with the viral vectors described above. 100,000 cells were plated in 12-well plates treated with rectronectin-fibronectin fragment (Takara) and treated with viral supernatant followed by spin-fection at room temperature at 1300xg for 45 minutes. Following two days of incubation at 37°C and 5% CO_2_, 1ug/mL of puromycin (Invivogen) was added to cell culture media to select for clones containing IRF7. Initial titration showed that a MOI of 2 was sufficient for stable expression of IRF7 and all cell lines generated for this study utilized an MOI of 2 for transduction.

A549 IRF7 deficient cells were transduced with the viral vectors generated above. 100,000 cells were plated in 12-well plates treated with rectronectin-fibronectin fragment (Takara) and treated with viral supernatant followed by spin-fection at room temperature at 1300xg. Following two days of incubation at 37°C and 5% CO_2_, cells were isolated and diluted for monoclonal isolation. Briefly, cells were diluted to a concentration of 0.3 cell/100 ul of media and plated in multiple 96-well plates and allowed to sit. Wells were then scanned 10-14 days later for colonies emerging from a single clone. Additional clones were validated using ELISA (PBL Assay Science, as described above) for IFN-α detection.

### RT-qPCR

RNA was extracted using the RNeasy mini kit (QIAGEN) and 1ug of RNA was used to make cDNA using SuperScript II ReverseTranscriptase (Thermo Fisher Scientific) according to the manufacturers’ protocol. cDNA was then diluted at a 1:20 ratio and qPCR was performed on a QuantStudio with iTaq Universal SYBR Green Supermix (Bio-rad) with the following forward and reverse primers.

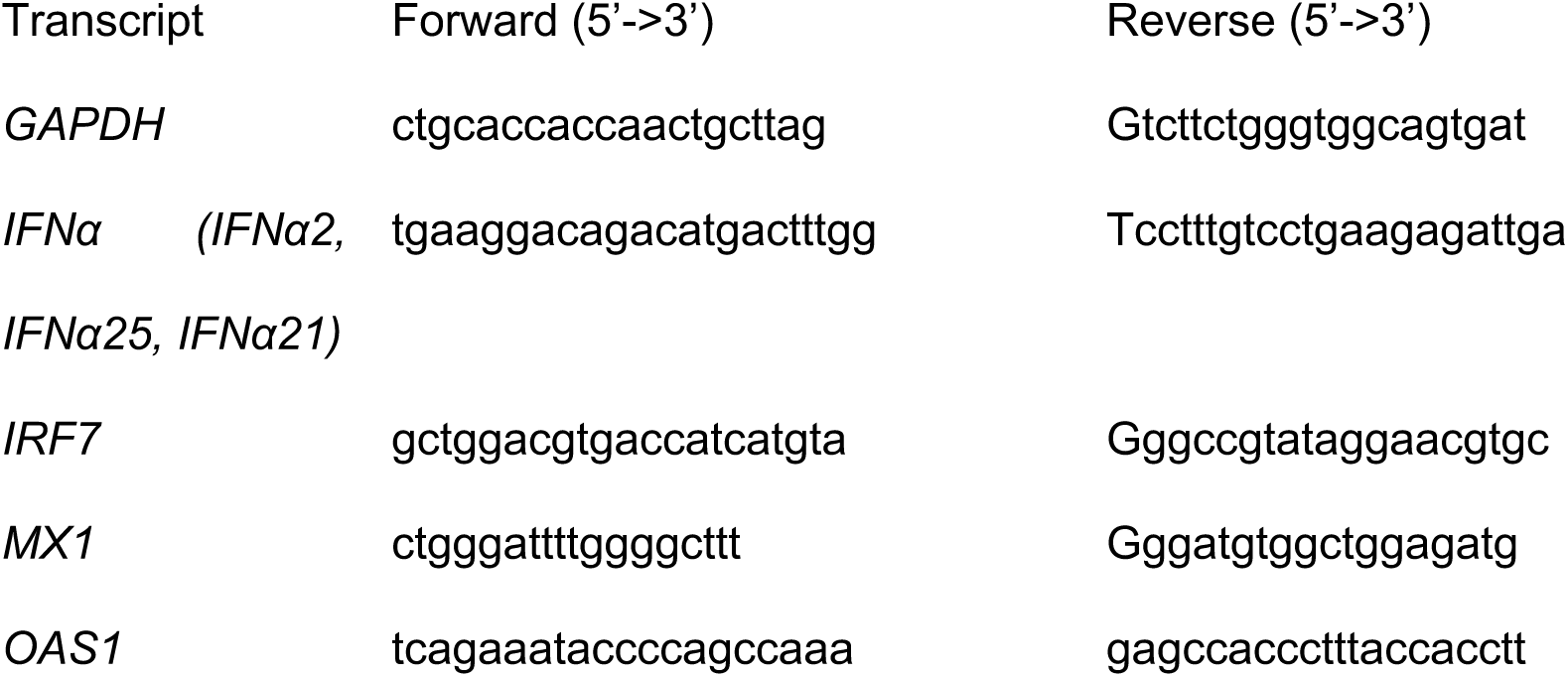

### Chemiluminescent Western Blot For pIRF7 and Total IRF7

Nuclear lysates of samples were mixed with loading buffer and DTT and heated at 95°C for 2 min. After cooling, the samples were loaded into a 4–12% Nu page Bis–Tris Gel (Invitrogen) and run for 90 min at 130 V in MOPS buffer, or 65 min at 130v in MES buffer. The gel was then transferred to a nitrocelulose membrane using the iBlot Machine (ThermoFisher Scientific). Samples were blocked in 5% milk in TBST (50 ml TBST plus 2.5 g dehydrated milk powder) for at least 1 h. The primary antibody (pIRF7: Cell Signaling) was diluted in 5% milk plus TBST at a 1:1000 ratio and the membranes incubated in primary antibody overnight at 4 °C with rocking. Membranes were washed in a TBST solution 3 times for 5 min each and were incubated in HRP conjugated secondary antibody in 5% milk in TBST for 1 h at room temperature with gentle shaking. Membranes were washed again in a TBST solution 3 times for 5 min each. Then, 2 ml of SuperSignal West Femto Maximum Sensitivity Substrate (Thermo Fisher Scientific) was added to each blot and incubated for 2 min. Substrate was removed before being imaged with the ChemiDoc Touch Imaging System (Bio-Rad).

### RNA-sequencing

We perform RNA-seq from experiments in both HEK and THP-1 cells. The mirVana RNA (Thermo Fisher Scientific) isolation kit was used to isolate total RNA from 6-well plates of 700,000 cells per well of each cell line in triplicate (at greater or equal to 90% viability) following the manufacturer’s recommended procedures. Libraries were prepared from RNA samples using the TruSeq Stranded Total RNA with Ribo-Zero Globin (Illumina, Inc., San Diego, CA) and sequenced at 150 paired end bases (50 M reads per sample) at the CCHMC Genomics Sequencing Facility.

RNA-seq datasets were processed using the nf-core pipeline ^98,99^ (v. 3.11.1) (https://nf-co.re/rnaseq). In brief, the samples underwent QC using FastQC v. 0.11.9 (https://www.bioinformatics.babraham.ac.uk/projects/fastqc/) and adaptors were trimmed and filtered using Cutadapt (v. 3.4) (https://cutadapt.readthedocs.io/en/stable/) ^100^. Other tools, such as RSeQC ^101,102^, Qualimap ^103,104^, dupRadar ^105^, and preseq (https://github.com/smithlabcode/preseq), were used to assess the quality of the datasets. Sequencing data were aligned to the hg19 genome using STAR (v. 2.7.9a). Ribosomal RNA was removed from the aligned datasets using the Silva Library (https://www.arb-silva.de/) and SortMeRNAM v. 4.3.4 (https://bioinfo.lif.fr/RNA/sortmerna/) to eliminate potential contamination from non-target species. Transcript abundance estimates were calculated using the GENCODE (v. 19) ^106^ annotation for hg19. Normalized gene read counts were calculated using DESeq2 ^107^. Transcript abundance estimates were calculated using GENCODE (hg19; v19)^106^ annotations with Salmon ^108^ (v1.10.1). The quantifications were imported into R (v4.1.3) and summarized at the gene level using tximport (v1.22.0) ^109^. Differential analyses were performed using DESeq2 ^107^ (v1.34.0) and genes were considered to be differentially expressed if the adjusted p-value was less than 0.05 and the absolute value of the log_2_ fold change was greater than 0.585 (i.e., a 1.5-fold linear change).

All datasets passed standard QC procedures and showed strong agreement between replicates. RNA-seq-based gene pathway enrichment analysis was performed with Enrichr ^110^ and gene sets from Reactome 2022 ^111^.

### Nuclear Extraction for Protein Binding Microarrays

50 million THP-1 cells stably expressing the lupus and protective haplotypes with a HA epitope tag (generated as described above) were maintained in T175 flasks prior to transfection with Poly(I:C). Three hours following 2.5 ug/mL Poly(I:C) transfection (see above), cells were centrifuged and washed in 1X PBS+ 1X protease inhibitor and centrifuged to remove PBS. Cell pellets were resuspended in Buffer A (10mM HEPES pH 7.9, 1.5 mM MgCl_2,_ 10mM KCl, 0.1 mM protease inhibitor cocktail, and 0.5mM of DTT) and incubated on ice for 10 minutes. Following incubation, 10% lgepal detergent was added and cell pellets were vortexed to ensure the detergent was spread throughout the sample. Following cell lysis, samples were centrifuged to pellet nuclei and cytosolic fractions were collected, snap frozen in liquid nitrogen, and stored at –80 °C. The nuclear pellet was washed with additional Buffer A. Pelleted nuclei were then resuspended in 4 aliquots of 45 ul of Buffer C per tube (20mM HEPES pH 7.9, 25% glycerol, 1.5mM MgCl_2_, 0.2 mM EDTA, 420 mM NaCl, 0.1mM protease inhibitor cocktail, and 0.5 mM DTT) and vortexed briefly followed by incubation with rotation and agitation at 4C for 1 hour. Following incubation, nuclei were pelleted and the nuclear extract was isolated, snap frozen with liquid nitrogen, and stored at –80 °C until application for the protein binding microarray.

### Protein Binding Microarray Experiments

#### Microarray design

The microarrays used for the PBM experiments were described previously in Andrilenas et al. ^55^. The 10,410 synthetic, ISRE-type sequences (**Figure 5)** were based on 2– and 3-bp-spacer ISREs defined by 5’-GANA-3’ type core elements separated by 2 or 3 bases, respectively. The microarray contained 6,594 2-bp ISRE variants based on 76 seed ISRE sites and all single-nucleotide variants of those seeds, as well as 3,456 3-bp ISRE variants based on 36 seed sites and all single-nucleotide variants.

#### PBM experiments

The microarray double-stranding procedure and nuclear extract PBM experiments were performed as described previously ^112,113^. Briefly, double-stranded microarrays were pre-wetted in HBS (20 mM HEPES, 150 mM NaCl) containing 0.01% Triton X-100 for 5 min and then de-wetted in an HBS bath. Next, the array was incubated with 180 uL of protein sample mixture: 45 uL of nuclear extract sample, 135 uL of binding reaction buffer [20 mM HEPES, pH 7.9, 1 mM DTT, 0.2 mg/ml bovine serum albumin, 0.02% Triton X-100, 0.4 mg/ml salmon testes DNA]. Note that the final salt concentration was ∼105 mM NaCl due to the high-salt nuclear extract sample. The protein was left to bind the microarray for 1 hour in the dark. The array was then rinsed in an HBS bath containing 0.05% Tween 20 and subsequently de-wetted in an HBS bath. After the protein incubation, the array was incubated for 20 min in the dark with 10 μg/ml primary antibody (Anti-HA tag antibody, ChIP-grade) in 2% milk in HBS. After the primary antibody incubation, the array was rinsed in an HBS bath containing 0.1% Tween 20 and de-wetted in an HBS bath. Microarrays were then incubated for 20 min with 20 μg/ml of Alexa 488-conjugated secondary antibody (Goat anti-Rabbit IgG (H+L) cross-adsorbed secondary antibody) in 2% milk in HBS. The array was rinsed in an HBS bath containing 0.05% Tween 20 and then placed in a Coplin jar containing 0.05% Tween 20 in HBS. The array was agitated in solution in a Coplin jar at 125 rpm on an orbital shaker for 3 min and then placed in a new Coplin jar with 0.05% Tween 20 in HBS to repeat the washing step. It was then placed in a Coplin jar containing HBS and washed for 2 min as described above. After the washes, the array was de-wetted in an HBS bath. Microarrays were scanned with a GenePix 4400A scanner and fluorescence was quantified using GenePix Pro 7.2. Exported fluorescence data were normalized with MicroArray LINEar Regression ^114^.

#### DNA-binding motif determination

Binding motifs were constructed using a seed and single-nucleotide variant approach described previously ^55,115^. The approach highlights the individual base preferences when the protein is binding to a specific seed element and allows direct comparison of the base preferences between distinct proteins at a common DNA element.

### Generation of Mutant Mice

C57BL/6 mice are homozygous for nucleotide sequences encoding an R at position 334 in IRF7 (the homologous position to human IRF7 412). CRISPR-Cas9 genome-editing with homologous recombination was used to produce heterozygous R/Q 334 mice from the C57BL/6 background. Mice were genotyped using Sanger sequencing with DNA from tail clips of the mice. RR (protective) and QQ (risk) mice were bred for experiments presented in Figure 7. Mice were bred and maintained in the vivarium adhering to IACUC guidelines.

### Epicutaneous Application of R848

Eight-week-old female RR and QQ mice were sensitized by epicutaneous application of 100 micrograms of R848 three times per week for 8 weeks. Serum samples were collected before and after 8 weeks of the treatment and tested for autoantibodies against double-stranded (ds) DNA using ELISA.

### Anti-double-stranded DNA (ds-DNA) ELISA

Plates were coated with 6 μg/mL ds-DNA in DNA Coating solution, washed with Tris-buffered saline (TBS) containing 0.05% Tween 20, and blocked with 1% BSA at 4ᵒC in 96-well plates. Following incubation, plates were washed and serum samples were incubated for an hour at room temperature on a plate shaker. Plates were washed and goat anti-mouse IgG HRP (horse radish peroxidase) was incubated for an hour. Plates were washed and TMB substrate was added, and absorbances for each well were read at 450 nm on a Biotek plate reader. Absorbance values were graphed using Graphpad Prism.

### Vesicular Stomatitis Virus (VSV) Infection

Eight-week-old male mice were intranasally infected with 1×10^6^ plaque forming units per mouse of the New Jersey serotype of VSV. Viral stocks were generated from a progenitor provided by Dave Masopust via propagation in BHK cells. Viral titers were determined using Vero cells. Mice were euthanized after 24 hours. Lung homogenates were prepared using a gentle tissue homogenizer and were stored at –80°C. Homogenates were aliquoted in small tubes to avoid frequent freeze-thaw cycles.

### Viral Plaque Assay

Vero cells were cultured and maintained in Dulbecco’s Modified Eagle Medium (DMEM). To titer virus, single cell suspensions of Vero cells were prepared using 0.25% trypsin and 0.2 million cells were seeded per well in 6-well plates. After overnight culture of cells, lung homogenates were thawed on ice and 10-fold serial dilutions were prepared. A range of sequential 10-fold dilutions were added to individual wells of Vero monolayers, followed by a 90 minute incubation and then overlay of cells with 0.5% SeaKem agarose in Eagle’s minimum essential medium (EMEM). Serially diluted aliquots of VSV stock were used as a positive control. To visualize the plaques, plates were stained with a second overlay of 0.5% agarose containing 1% neutral red on day 2. Plaques were counted within 24 hours of staining. Viral plaques were counted, whereafter wells with counts between 10 and 60 were used to quantity virus concentration in the original volume of lung homogenate using the following formula: (average number of plaques) x (overall dilution factor).

## STAR METHODS TABLE

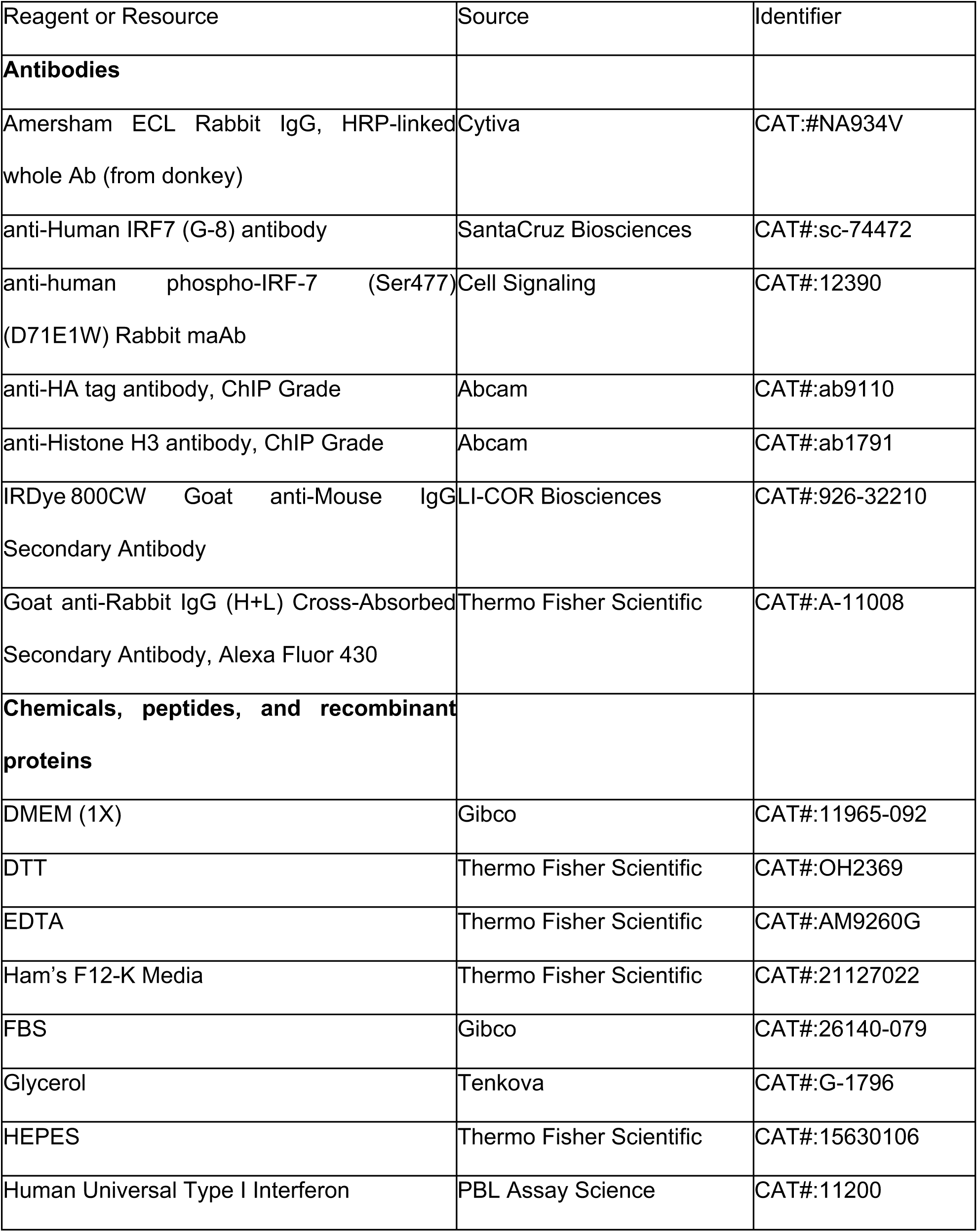
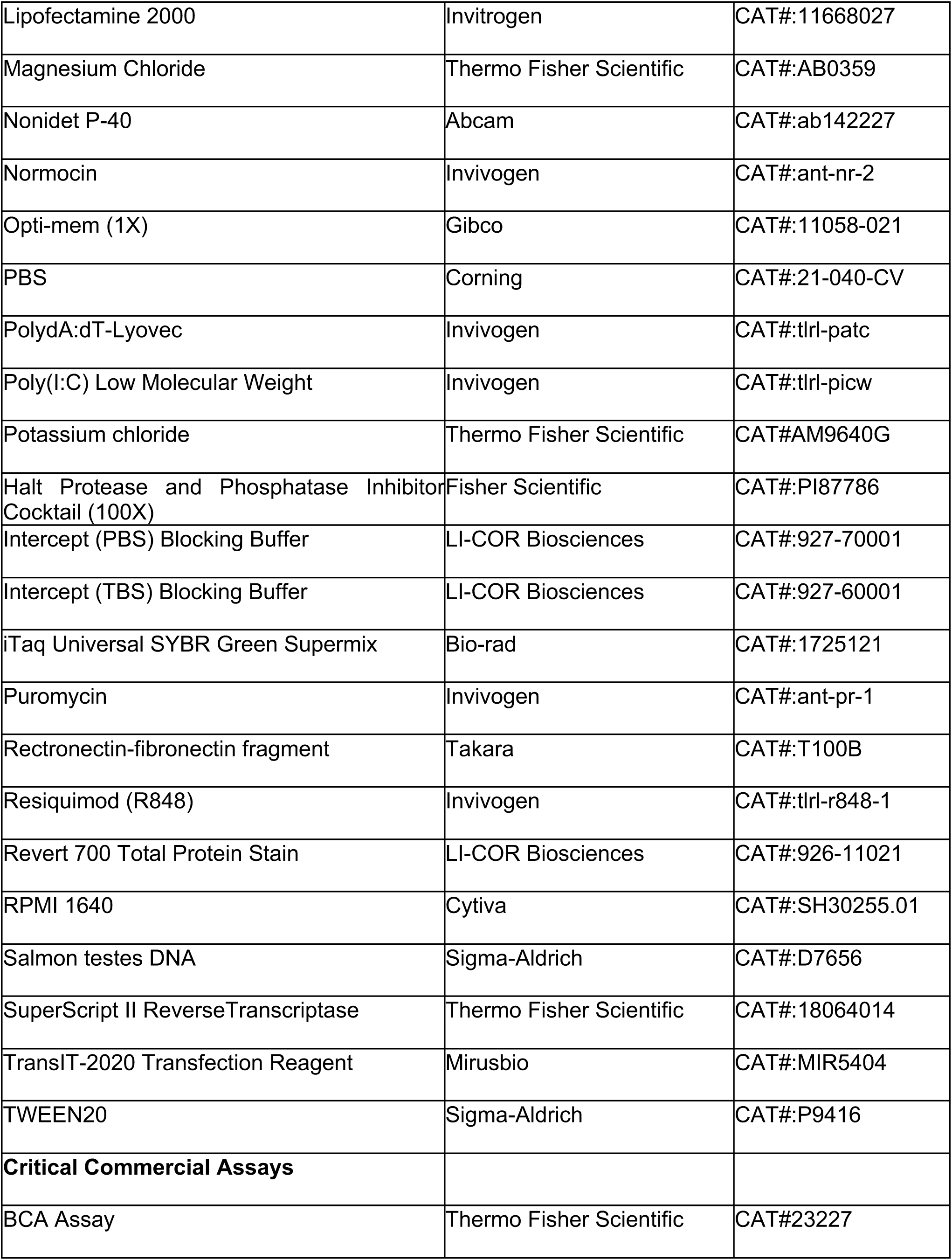
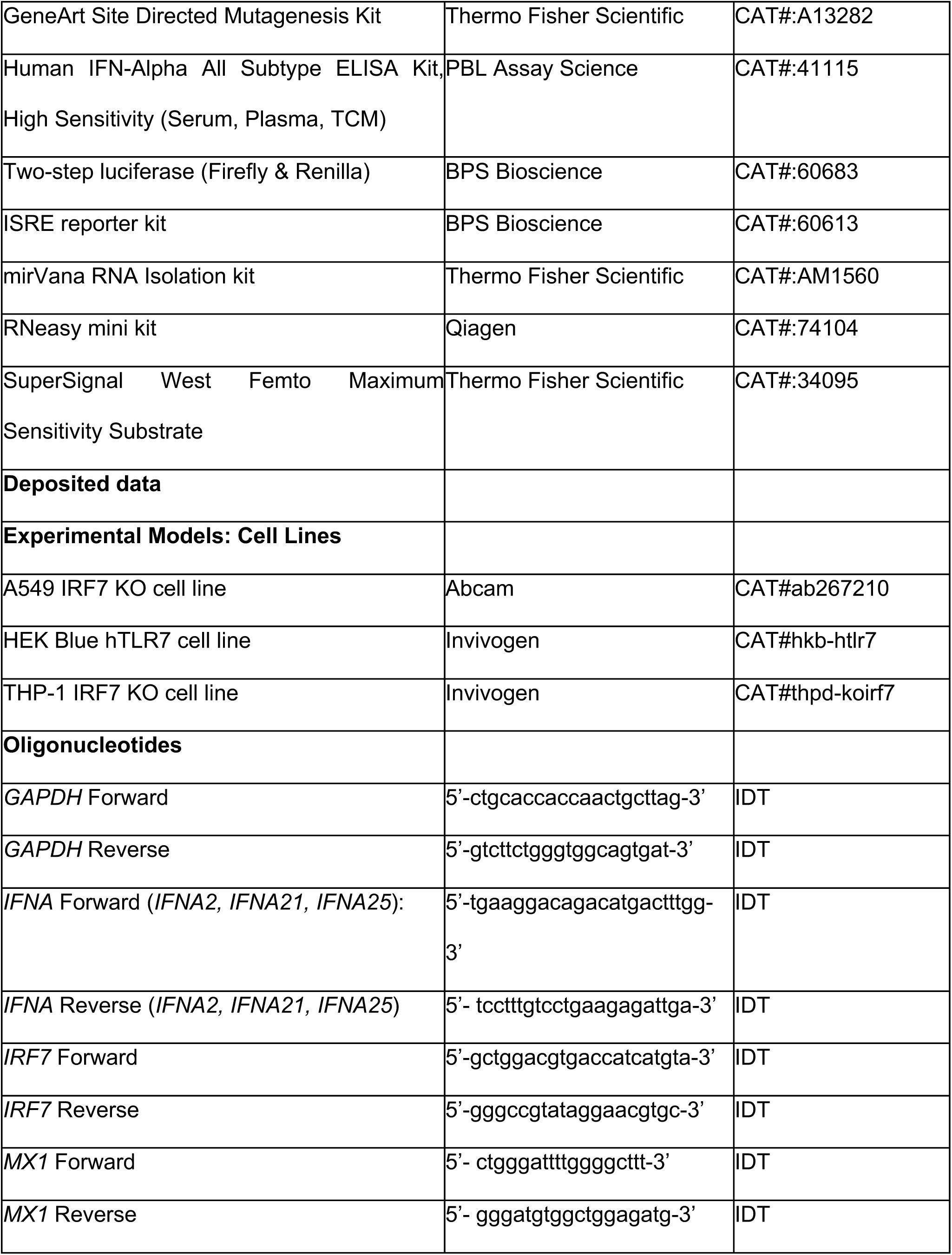
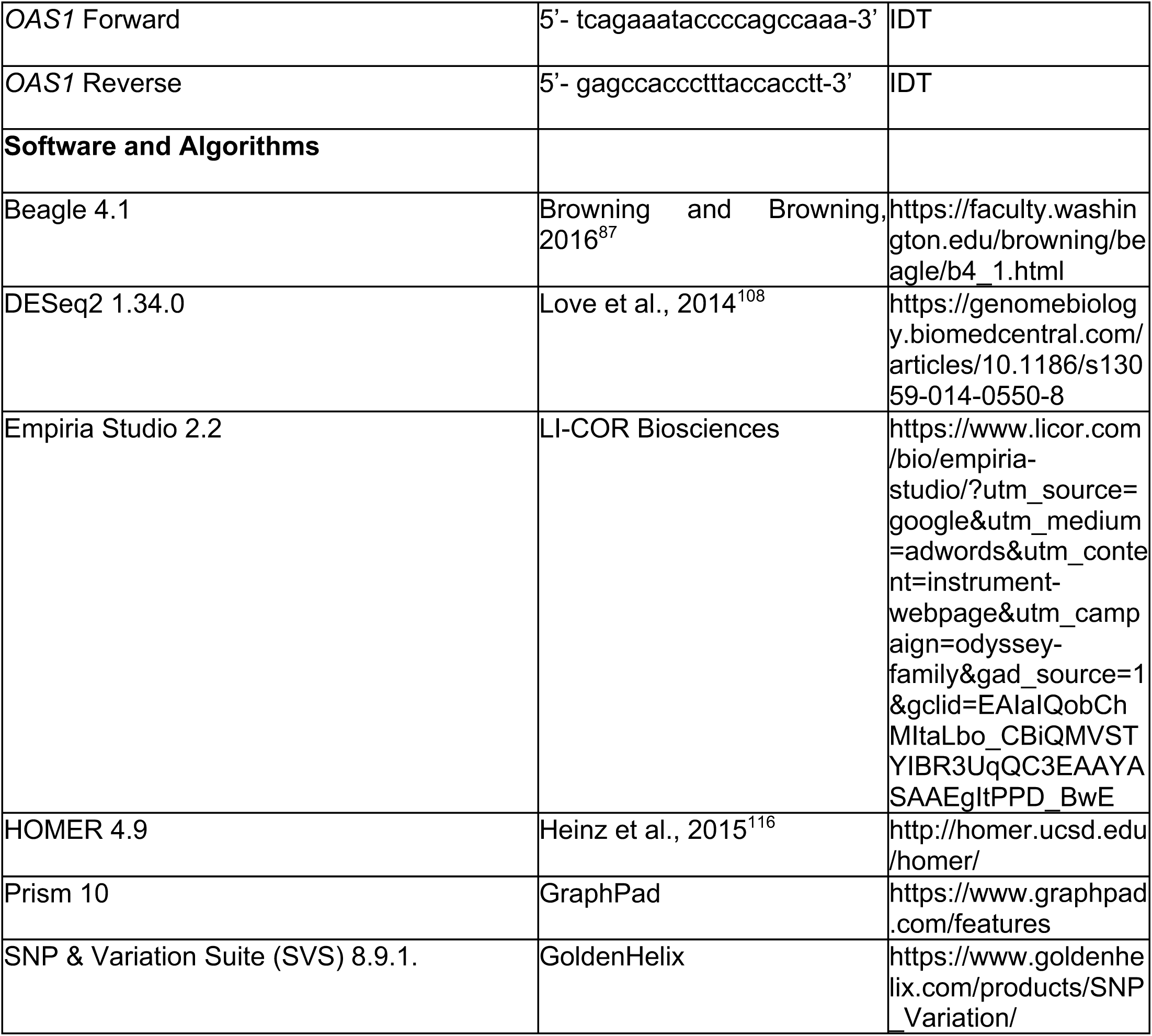

## SUPPLEMENTAL VIDEOS AND EXCEL TABLE TITLES AND LEGENDS

**Table S1**. Power calculations (a=0.05) to detect differences in allele frequencies of the Q412R haplotype in four ancestral groups in a cohort of 7,137 individuals with SLE and 6,329 individuals without SLE. Related to Figure 1.

**Table S2**. QC Summary Report for all RNA-seq datasets in HEK and THP-1 experiments. Related to Figure S3 and Figure 4.

**Table S3**. Gene expression (TPM) information for each cell line. Related to Figure 4.

**Table S4**. Differential expression results from DESeq2 (see Methods) for the lupus risk versus protective haplotype RNA-seq experiments in THP-1 and HEK Blue hTLR7 cell lines. Related to Figure 4.

**Table S5**. Pathway Enrichment (Reactome 2022) results for differentially expressed genes between the risk and protective IRF7 haplotypes using ENRICHR. Related to Figure 4.

## ABBREVIATIONS

cGAS: Cyclic GMP-AMP synthase
ELISA: Enzyme-linked immunosorbent assay
GFP: Green fluorescent protein
GWAS: Genome-wide association study
HEK293: Human embryonic kidney cell line
IFN-I: Type I interferon
IFNAR: Type I interferon receptor
IAV: Influenza virus A
IRF: Interferon regulatory factor
ISG: Interferon-stimulated gene
ISRE: Interferon-stimulated response element
KO: Knockout
LD: Linkage disequilibrium
PBM: Protein binding microarray
PRR: Pattern recognition receptor
R848: Resiquimod
RIG-I: Retinoic acid-inducible gene-1
SLE: Systemic lupus erythematosus
STING: Stimulator of interferon genes
TLR: Toll-like receptor
TSS: Transcription start site
VSV: Vesicular stomatitis virus

