## Supplementary material for "A highly prevalent lupus risk haplotype increases IRF7-dependent induction of IFN-α, enhancing antiviral defense and exacerbating autoimmunity": Table S1

| Ancestry | # of lupus Cases | # of controls | Frequency of Risk Allele in Cases | Frequency of Risk Allele in Controls | Power to Detect Differences |
|---|---|---|---|---|---|
| European | 3615 | 3150 | 77.0% | 73.0% | 96.7% |
| African | 1458 | 1673 | 54.0% | 49.0% | 79.8% |
| East Asian | 1202 | 1205 | 98.0% | 97.1% | 30.7% |
| Central and Latin American Ancestry | 862 | 301 | 70.0% | 67.1% | 15.6% |

Supplemental Table 1. Power calculations (a=0.05) to detect differences in allele frequencies of Q412R haplotype in four ancestral groups in a cohort of 7137 individuals with SLE and 6329 individuals without SLE.
