## Supplementary figures and images for "A highly prevalent lupus risk haplotype increases IRF7-dependent induction of IFN-α, enhancing antiviral defense and exacerbating autoimmunity"

### Figure S1

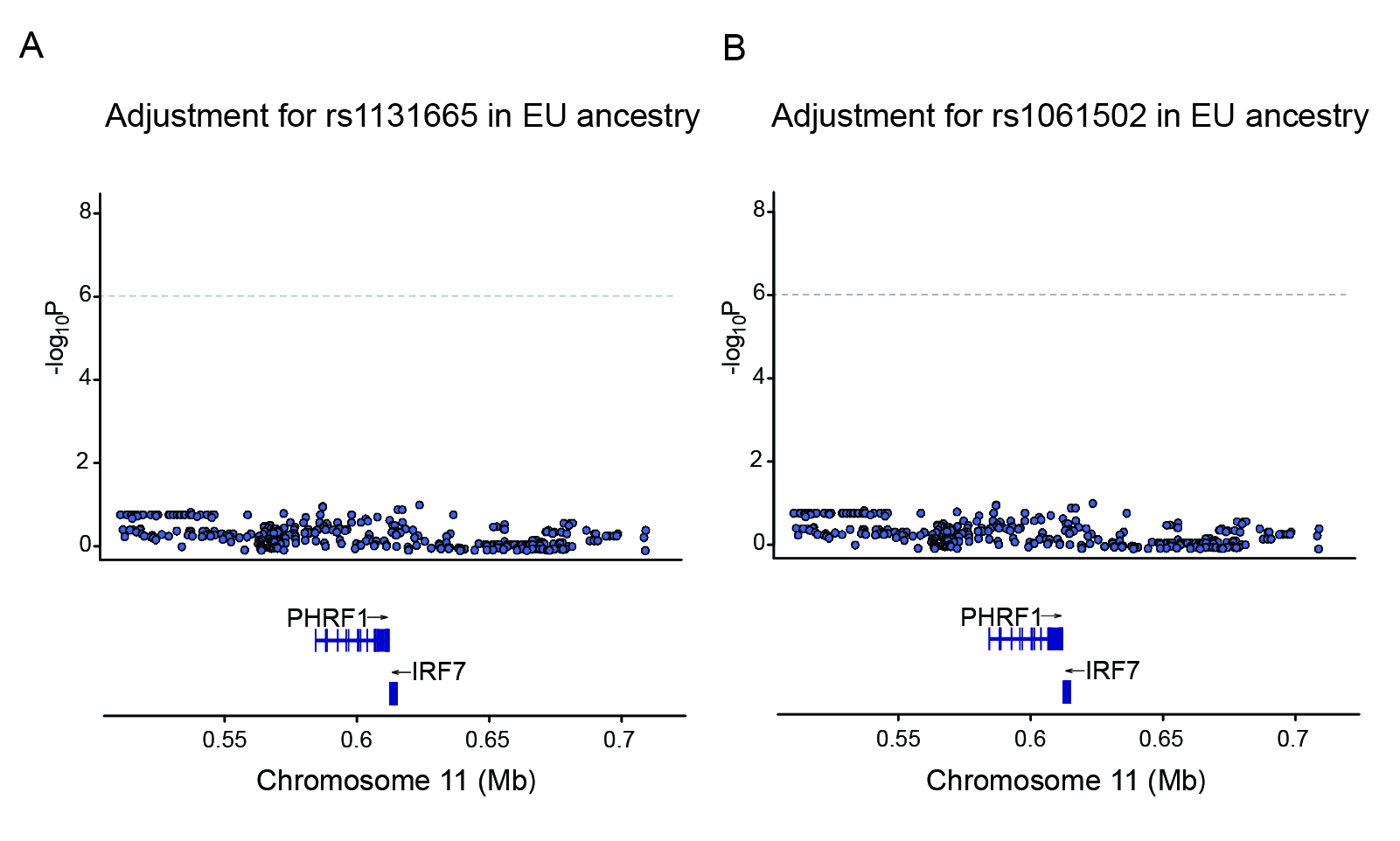

### Figure S2

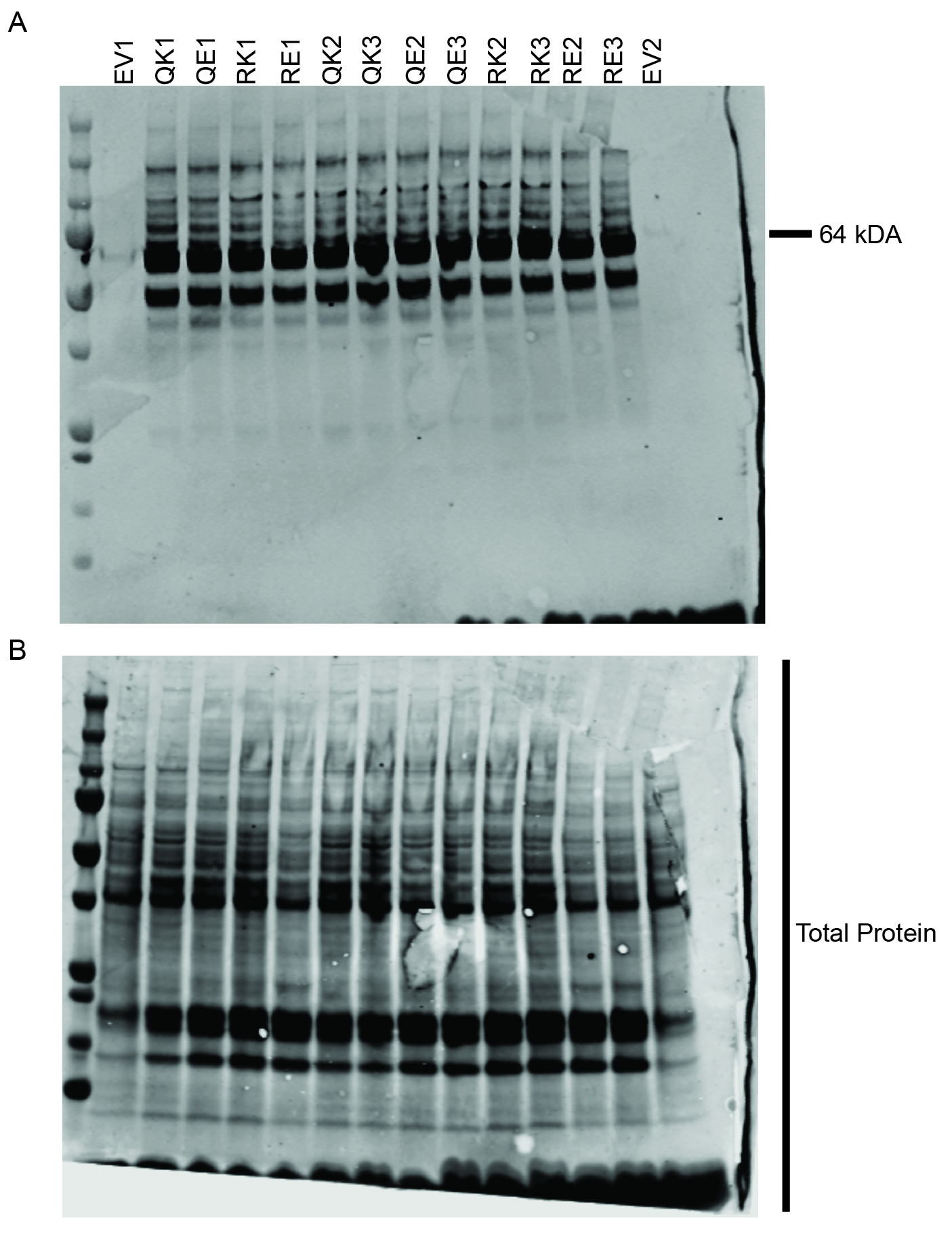

### Figure S3

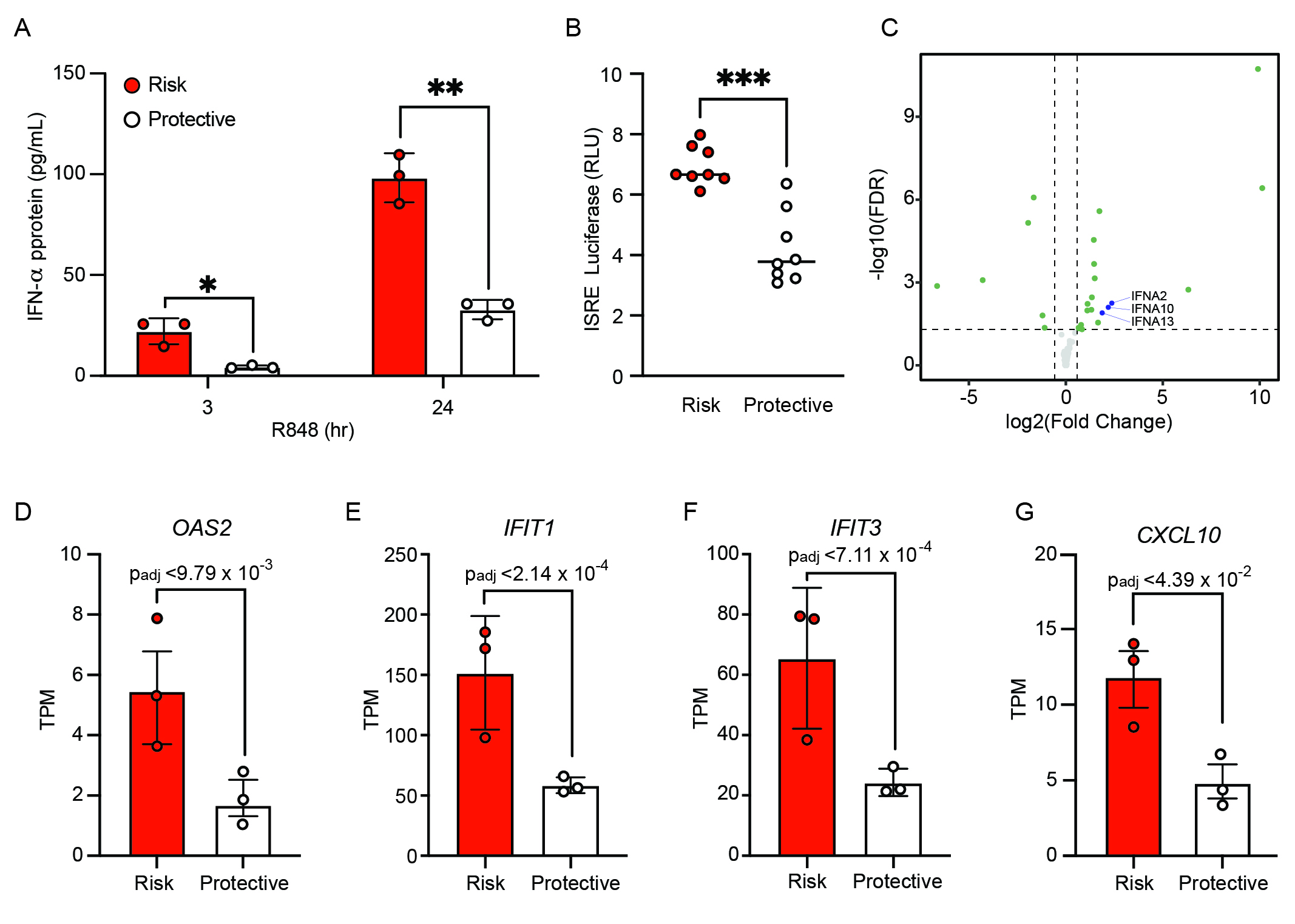

### Figure S4

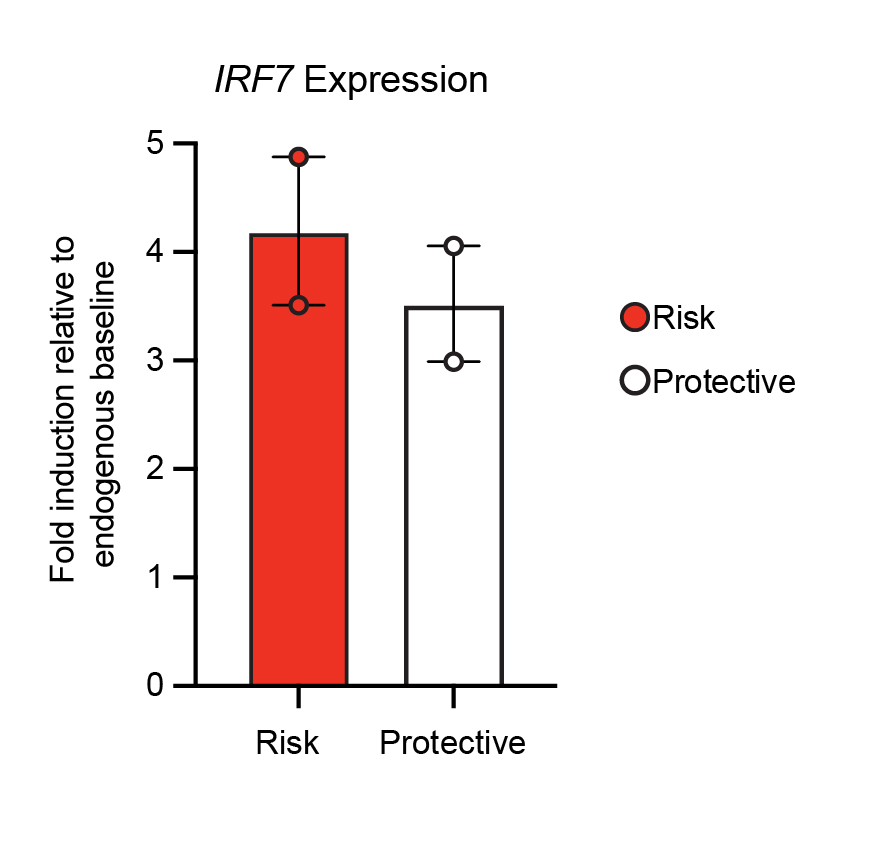

### Figure S5

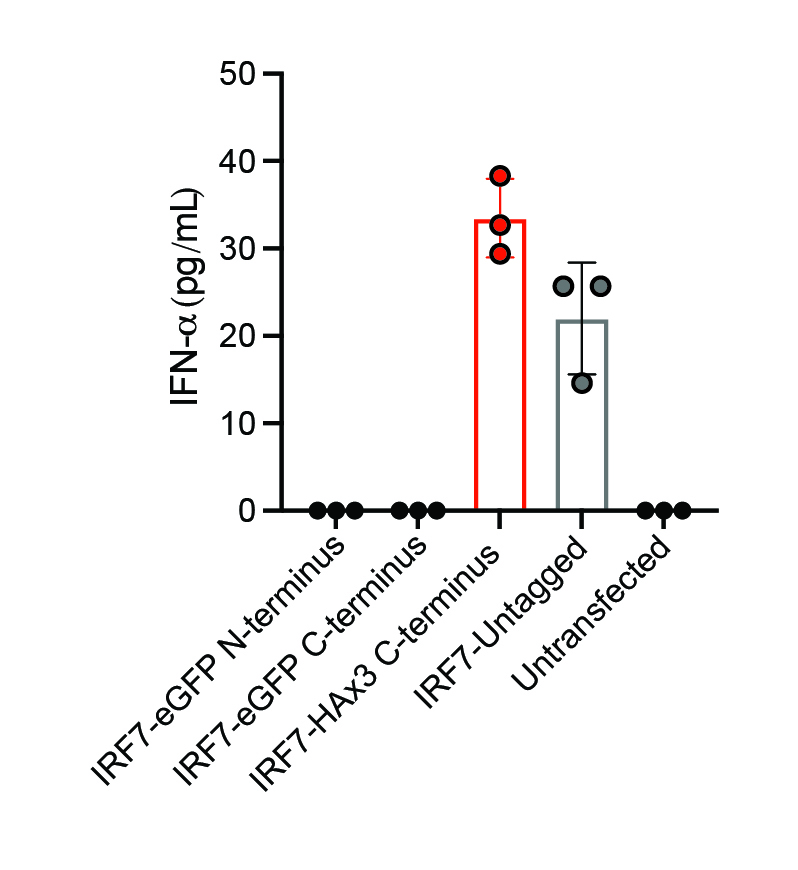

### Figure S6

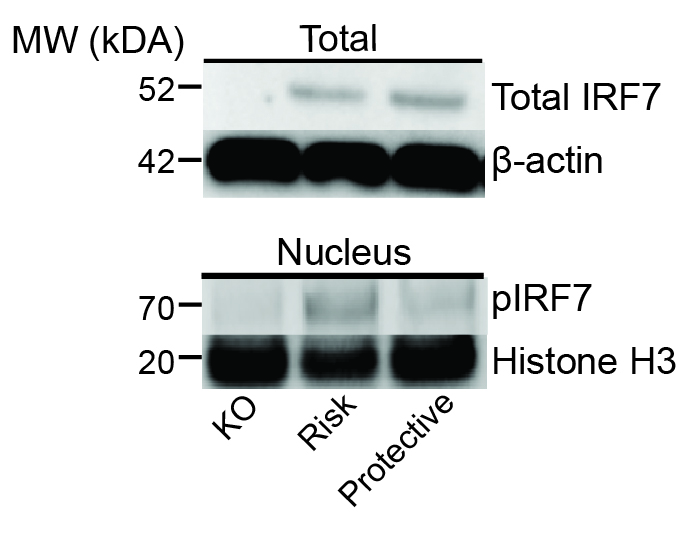

### Figure S7

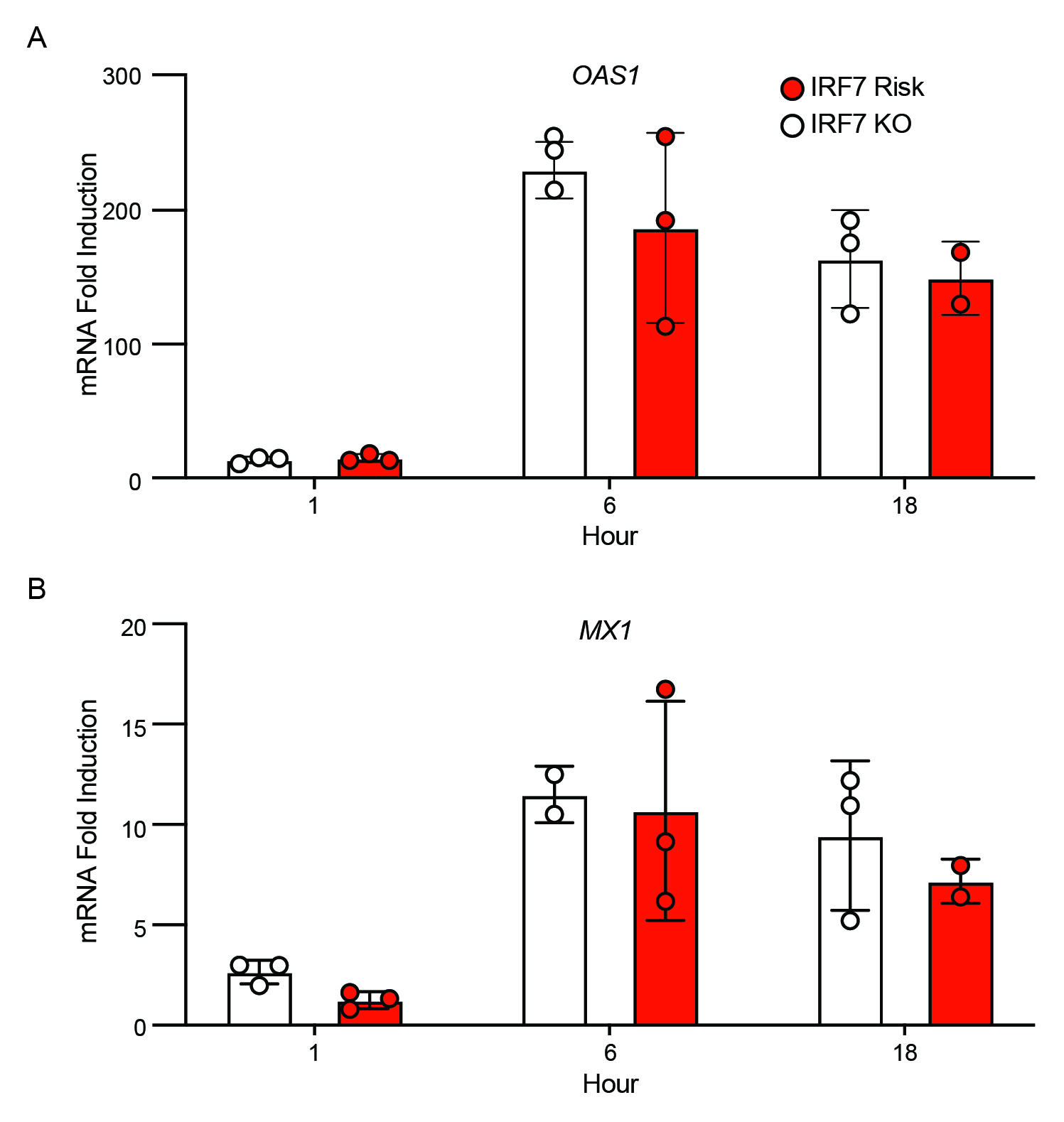

### Figure S8

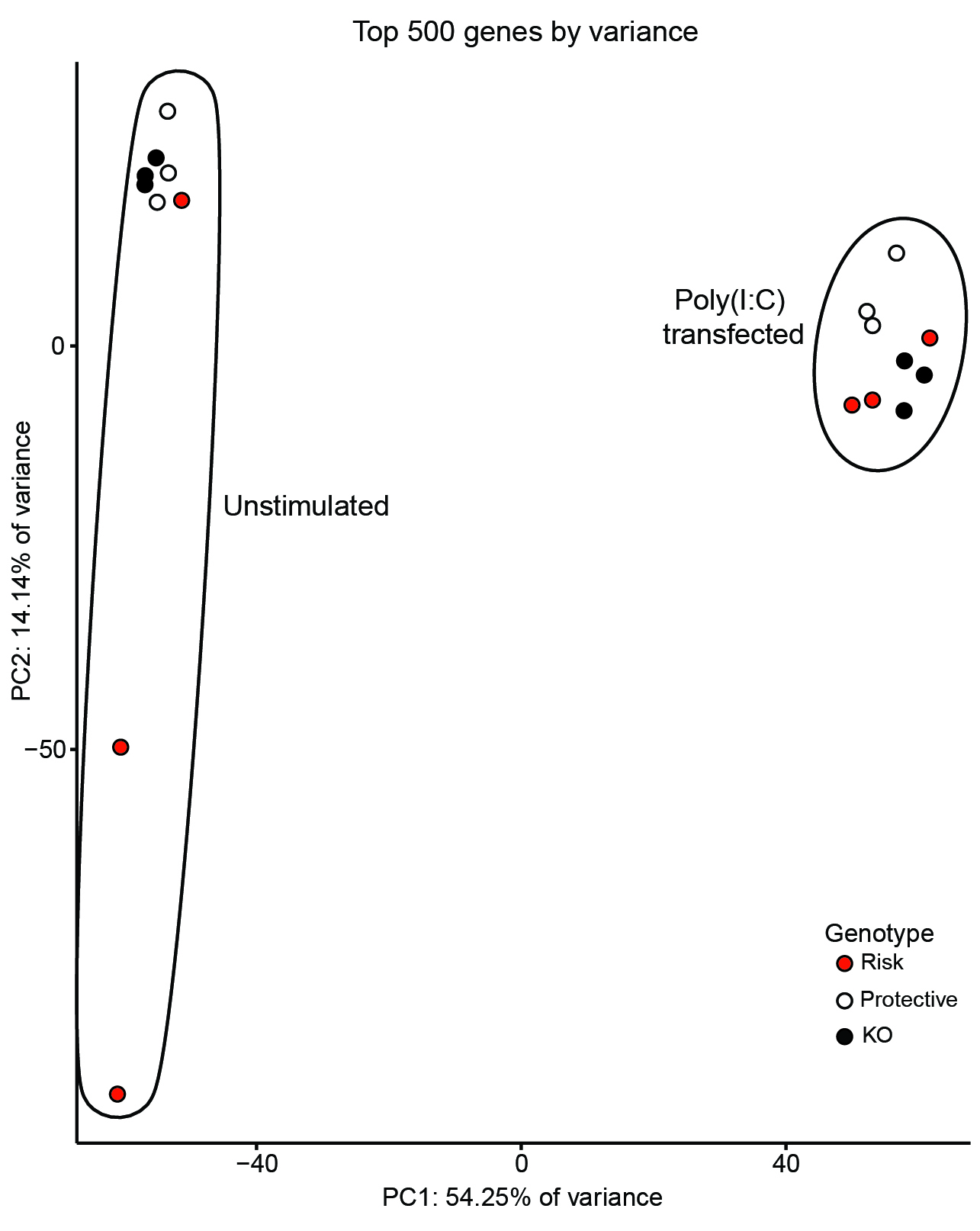

### Figure S9

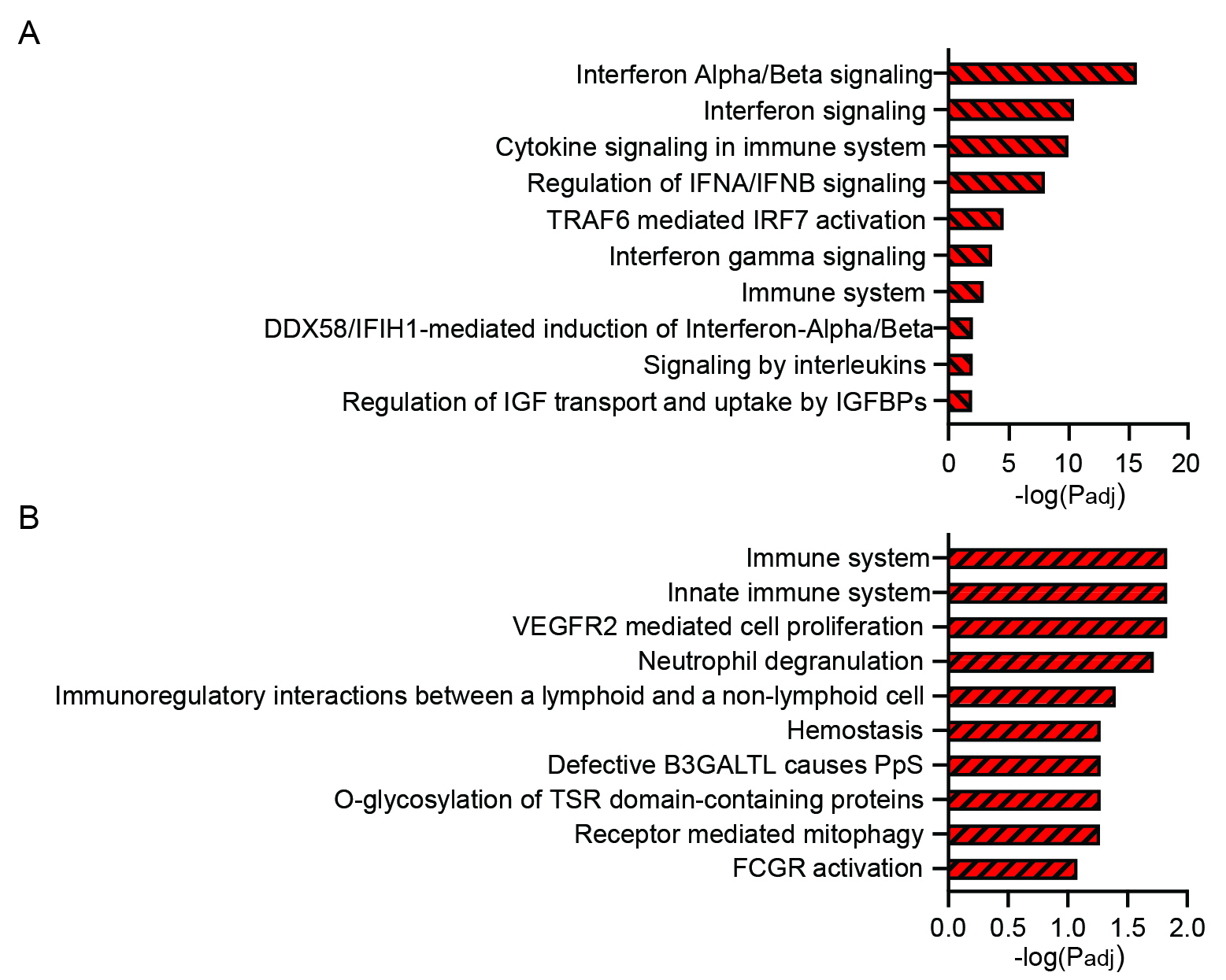

### Figure S10

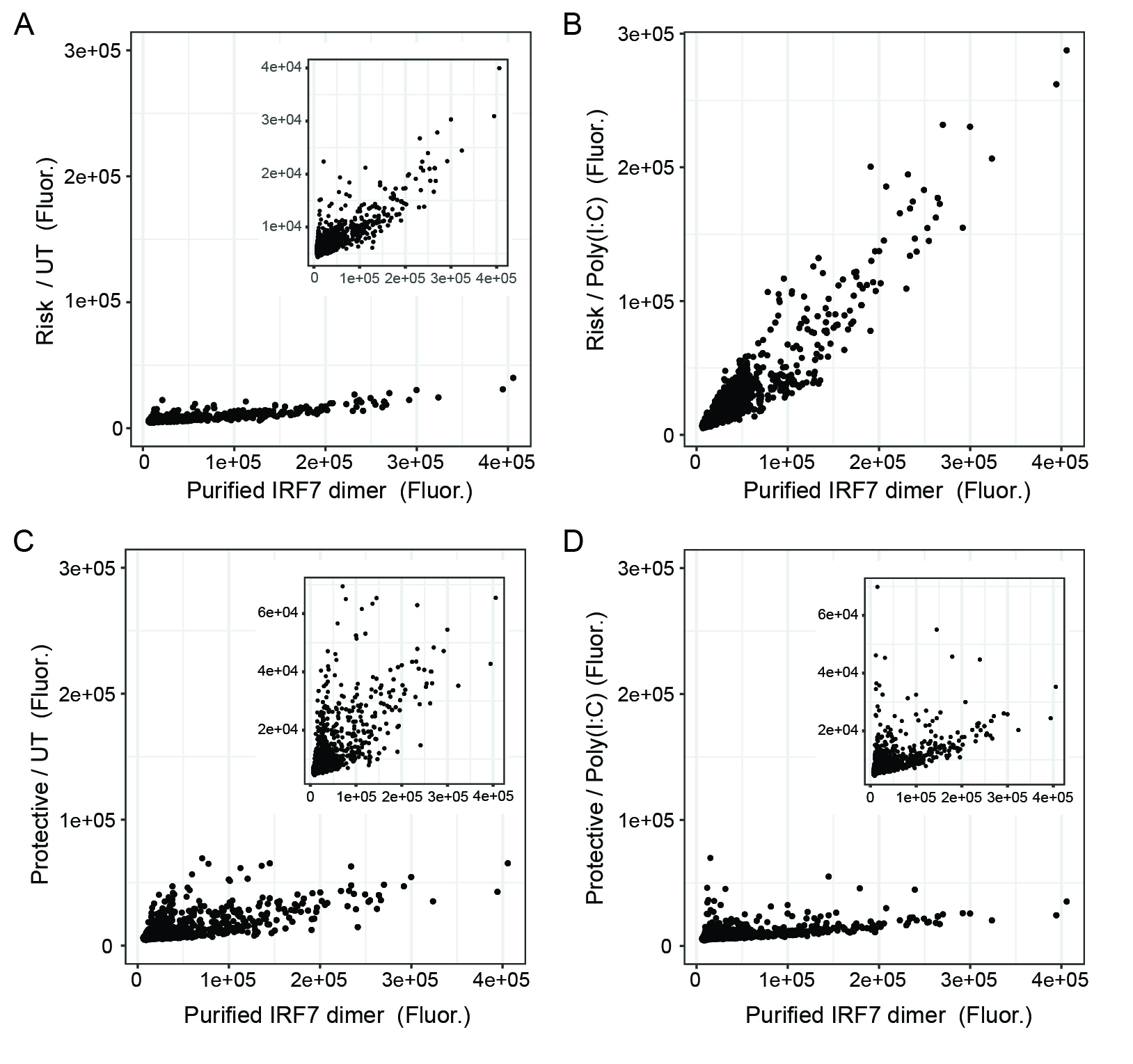

### Figure S11

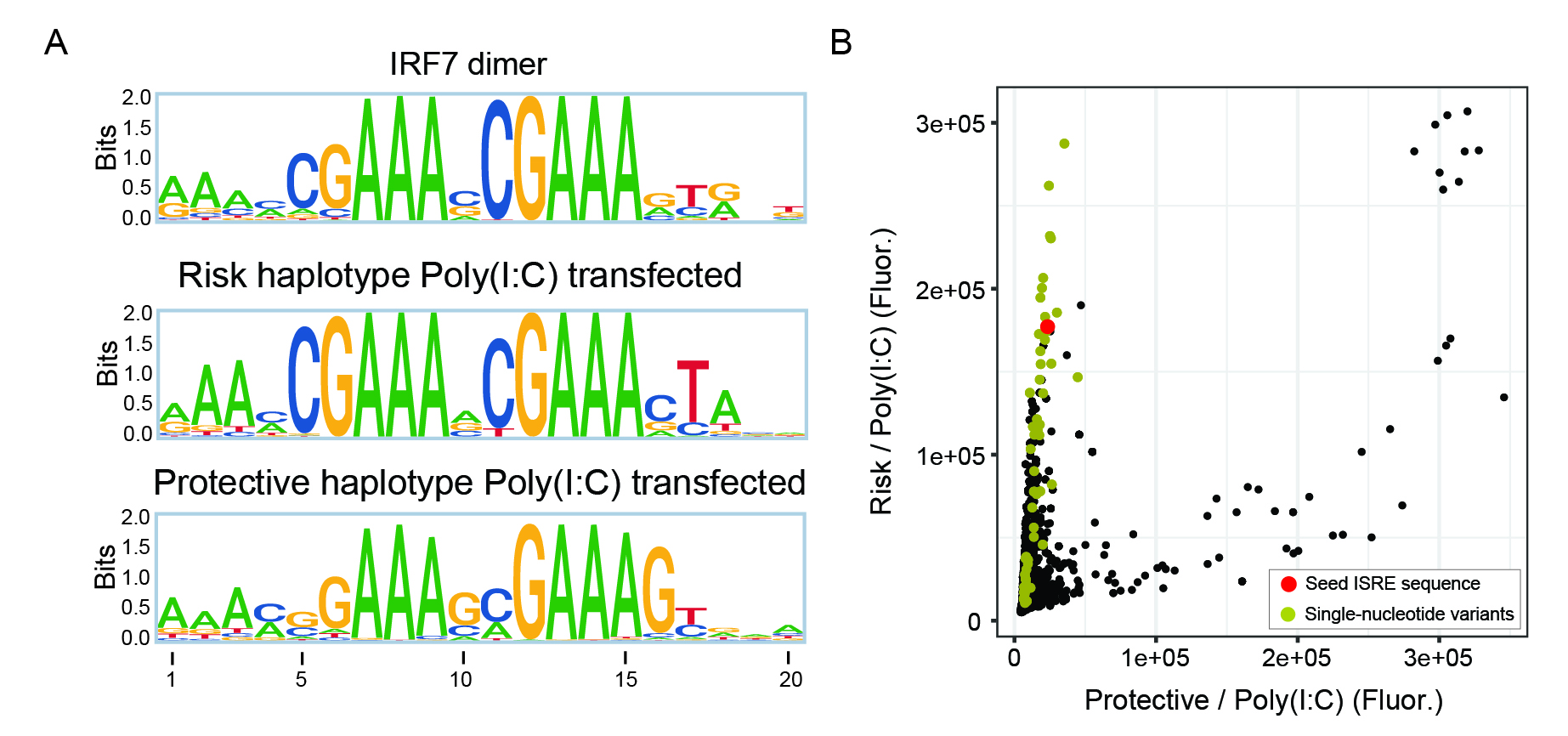

### Figure S12

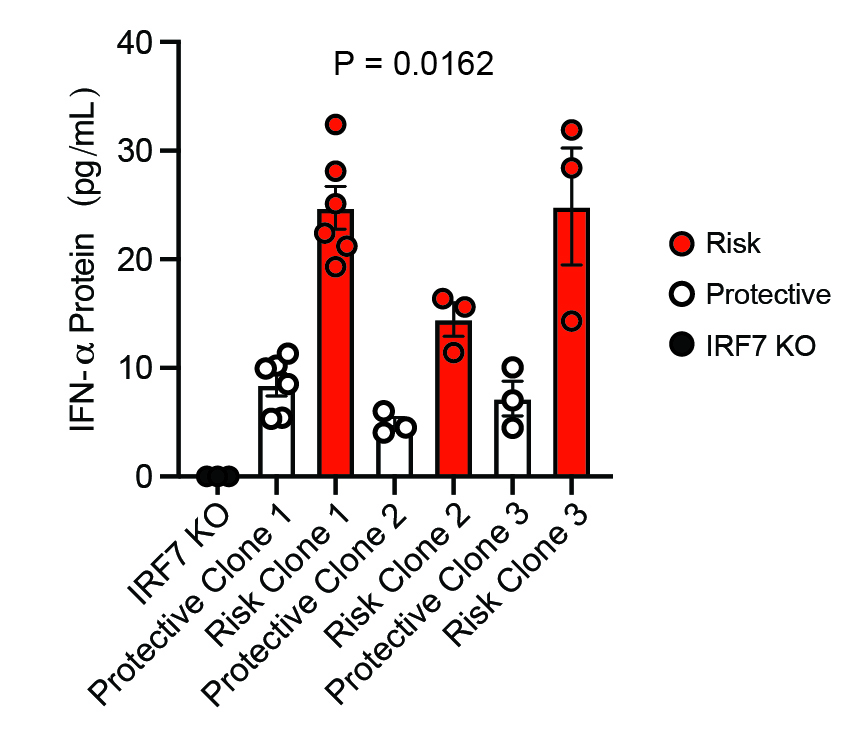
